# Impaired Reward-Based Learning but Preserved Motor Invigoration in Chronic Stroke

**DOI:** 10.1101/2025.10.31.25338954

**Authors:** S. Sporn, M. Herrojo Ruiz, R. Fathana, C. Zich, S. Bestmann, N. S. Ward

**Affiliations:** Department of Clinical and Movement Neuroscience, Queens Square Institute of Neurology, UCL, UK; Department of Psychology, Goldsmiths, University of London, UK; Wellcome Centre for Integrative Neuroimaging, Nuffield Department of Clinical Neurosciences, University of Oxford, UK; Medical Research Council Brain Network Dynamics Unit, University of Oxford, UK; Department of Imaging Neuroscience, Queens Square Institute of Neurology, UCL, UK; Centre for Clinical Neuroscience, Hospital Universitario Los Madroños, Brunete, 28690, Spain

**Keywords:** Stroke, reward-based learning, computational modelling

## Abstract

Reward provides a feedback signal that modulates behaviour through several mechanisms, including invigorating performance and learning of action–outcome associations to guide future choices. After stroke, the ability to utilise reward feedback can be impaired, which may limit the benefits of rehabilitation approaches that use reinforcement. One possibility is that stroke causes a global impairment of reward processing, leading to both reduced invigoration and diminished learning from feedback. Alternatively, reward processing may be selectively disrupted, such that either invigoration or the ability to update beliefs from reward feedback is disproportionately affected.

To test these competing hypotheses, we recruited forty chronic stroke survivors and thirty age-matched healthy controls to complete a probabilistic reversal learning task with both their strong (non-paretic/dominant) and weak (paretic/non-dominant) limb. On each trial, participants reached to one of two targets associated with different reward probabilities that changed unpredictably over time, requiring continued monitoring of outcomes and adaptation of choice behaviour.

Stroke survivors showed reduced reward-based learning compared to controls, expressed as lower overall choice accuracy and a greater tendency to switch responses after rewarded trials (i.e., lower win–stay rates), particularly when using the weak upper limb. Control analyses confirmed that these selective impairments were not explained by general motor impairment or cognitive deficits. To identify the putative computations underlying these behavioural differences in reward-based learning we used an established model of hierarchical Bayesian inference, the Hierarchical Gaussian Filter (HGF). The HGF characterises learning dynamics as trial-by-trial updating of an agent’s beliefs about action–outcome probabilities and their change over time (environmental volatility). Compared to healthy controls, stroke survivors were slower to update their beliefs about action–reward contingencies, an effect most pronounced for the weak upper limb, whereas updating beliefs about environmental volatility remained intact. Reward-based invigoration was also preserved: strong trial-by-trial predictions about action–reward contingencies were associated with faster movement times, with comparable slopes of this association across groups, indicating that motivational drive was maintained in patients despite overall slower performance.

This behavioural dissociation between preserved motivational invigoration but impaired probabilistic reward-based learning highlights a key translational opportunity: to leverage intact motivational pathways to enhance rehabilitation intensity and compliance, and to develop adaptive feedback strategies that compensate for impaired reward learning. Harnessing these complementary approaches could strengthen recovery outcomes and support greater long-term independence after stroke.

## Introduction

Stroke is the leading cause of neurological disability worldwide^1–3^. In the UK, stroke survivors are projected to rise by 62% to 2.1 million by 2035, with annual costs exceeding £75 billion nearly half the NHS budget. Over 173,000 working-age people will be lost from the workforce^4^. Upper limb impairment affects over 60% of survivors, causing long-term disability and severely limiting independence^5–7^. Improving recovery is therefore both a clinical and economic priority.^1–34^

Promoting recovery through rewards is one focus of rehabilitation^9^. Reward shapes behaviour through several mechanisms, including (1) invigorating performance, leading to increased movement speed and force^10–15^; and (2) promoting learning by strengthening action–outcome associations that guide decision-making and action selection^10,16–19^. These mechanisms rely on dopaminergic contributions^20,21^, with additional neurotransmitter systems—including serotonin and noradrenaline—also involved in motor vigour and belief updating, respectively^22,23^. However, whether these mechanisms remain intact after stroke is unclear.

Some studies report impairments in reward-based learning in both acute and chronic stroke^24–26^ and a reduced sensitivity to reward^27,28^, while others find preserved learning capacity^26^ and intact reward sensitivity^29^. Note here that the term *reward sensitivity* has been defined differently across studies. In some cases, it refers to changes in choice behaviour (i.e., win-stay rates) reflecting how effectively rewards reinforce future actions^24,25^. In others, it describes *invigoration*, where higher anticipated rewards accelerate movement initiation and execution^27–29^. Importantly, to date, no study has systematically assessed both mechanisms in stroke—reward-based learning and reward-driven invigoration—within the same individuals. This gap leaves open a critical question: is reward processing globally disrupted after stroke, selectively impaired, or largely preserved?

Understanding the nature of these changes is essential for developing effective neurorehabilitation strategies. If reward-based learning is preserved, it could be leveraged to promote long-term behavioural change; if invigoration remains intact, it could be used to boost compliance and training intensity—two key determinants of rehabilitation success^30^. Conversely, if either mechanism is compromised, tailored feedback strategies will be required. Such mechanistic understanding is critical for integrating reward-based principles into virtual reality, robotics, and AI-driven rehabilitation platforms that aim to deliver engaging, personalised home-based therapy at scale^9,31^.

Building on this rationale, the overall aim of this study was to determine whether chronic stroke survivors show impairments in reward-based learning and/or movement invigoration, and to identify which computational processes underlie these deficits. We define reward-based learning as the ability to infer action–outcome contingencies and their change over time (volatility), and invigoration as the tendency to move faster when holding strong expectations about reward probability^32^. We address three questions: First, do stroke survivors earn fewer rewards and show altered stay/shift behavioural strategies following rewarded and non-rewarded outcomes, with either their strong or weak upper limb? Second, are any such deficits accounted for by impaired belief updating about action–outcome contingencies and their change over time (environmental volatility)? Third, is the motivational effect of reward expectation on movement time (i.e., reward-based invigoration) altered after stroke?

To address these questions, we compared 40 chronic stroke survivors (SK) and 30 age-matched healthy controls (HC) on a probabilistic reward-based learning task with changing action–outcome contingencies^32,33^. Participants performed the task with both their weak (paretic/non-dominant) and strong (non-paretic/dominant) limb. On each trial, participants made a reaching movement to select between two targets, one of which was more likely to yield reward. These reward contingencies reversed unpredictably, requiring participants to integrate outcome feedback over time to update their beliefs about which target was currently more rewarding. This task captures key aspects of real-world decision-making under uncertainty and has been extensively validated in healthy populations and across neurological and psychiatric conditions (e.g., Parkinson’s disease^32^, mood disorders^34^), making it well-suited to assess whether stroke survivors can adaptively update beliefs about action-outcome mappings by integrating reward feedback.

To examine the computations underlying reward-based learning and belief updating in this task, we used the Hierarchical Gaussian Filter (HGF)^35,36^, a hierarchical Bayesian inference model that estimates how individuals learn changing action–outcome contingencies over time. The HGF provides a formal framework to explain observed decisions (responses) as a function of latent processes such as uncertainty estimation and belief updating. It builds on the notion that the brain maintains a hierarchical generative model of the environment and continuously updates beliefs in response to incoming information, including rewards^37^. Within this framework, belief updates are driven by prediction errors (PEs)—the discrepancy between expected and actual outcomes—and are modulated by precision weights, where precision is defined as the inverse variance (or uncertainty) of belief distributions. The HGF has proven useful for identifying computational mechanisms underlying altered learning in both clinical and subclinical conditions^32,33,38^.

By fitting the HGF to each participant’s trial-by-trial choices, we examined whether impairments in reward-based learning after stroke arise from altered updating of beliefs about action–reward contingencies, their volatility, or both. In addition, by deriving trial-wise trajectories of reward expectations for each participant from the model, we determined how these predictions influenced movement times, allowing us to assess potential changes in reward-based invigoration after stroke^32,34^. Together, these analyses allowed us to dissociate the effects of stroke on the reward-based learning process from those on the motivational drive to act.

## Methods

### Experimental Subjects

Forty chronic stroke patients (≥6 months post-onset) were recruited from the Queen Square Upper Limb (QSUL) Rehabilitation Programme and thirty age-matched healthy controls participated. Inclusion criteria for stroke patients were (1) first-ever stroke and (2) absence of other brain injury, neurological disorder, or major psychiatric illness. Exclusion criteria were (1) hemi-spatial neglect or hemianopia, (2) severe aphasia, and (3) pain limiting task performance or protocol adherence. Inclusion criteria for controls were (1) naïve to the task paradigm and (2) no self-reported neurological or psychiatric history. All participants provided written informed consent prior to participation. Clinical and demographic characteristics are summarised in **Table 1** (see **Table S1** for individual data and lesion site information). Because age data were not normally distributed, group differences were assessed using a Wilcoxon rank-sum test, with rank-biserial correlation (*r*) reported as the effect size. The test showed that (*p* = 0.136, *z* = –1.49, *r* = –0.14; small effect; BF = 0.33; substantial evidence for H_0_), the groups were not significantly different with regard to their age.

**Table 1.**
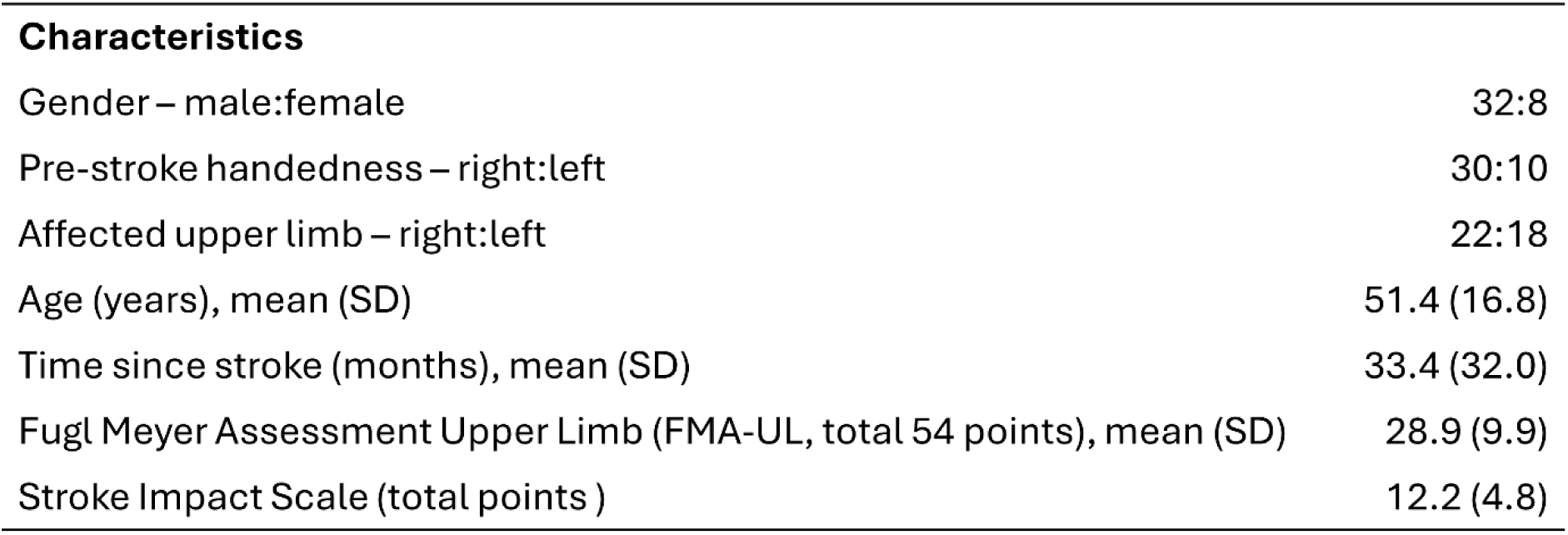
Clinical and demographic characteristics participants.

**Characteristics**
|  |  |
| --- | --- |
| Gender – male:female | 32:8 |
| Pre-stroke handedness – right:left | 30:10 |
| Affected upper limb – right:left | 22:18 |
| Age (years), mean (SD) | 51.4 (16.8) |
| Time since stroke (months), mean (SD) | 33.4 (32.0) |
| Fugl Meyer Assessment Upper Limb (FMA-UL, total 54 points), mean (SD) | 28.9 (9.9) |
| Stroke Impact Scale (total points) | 12.2 (4.8) |

### Ethical Approval

This study was reviewed and approved by the London Camden and Kings Cross Research Ethics Committee (REC reference 17/LO/1466) and conducted in accordance with the Declaration of Helsinki.

### Materials

The experiment was conducted using the KINARM Exoskeleton (BKIN Technologies Ltd, Kingston, ON, Canada; **Figure 1a**), a robotic device that records upper-limb kinematics during planar reaching tasks. The exoskeleton supports the participant’s arms and hands, constraining movements to a horizontal plane involving flexion and extension of the shoulder and elbow joints. Participants were seated with their arms abducted ∼85–90° from the trunk, each arm supported by a customised linkage for comfort and alignment. A 2D virtual reality display, integrated in the movement plane, provided visual targets and feedback. The system was calibrated before each session to ensure accurate limb–cursor alignment. The robot offered full gravitational support but no movement assistance. Further technical details are described elsewhere^39–41^.

**Figure 1.**
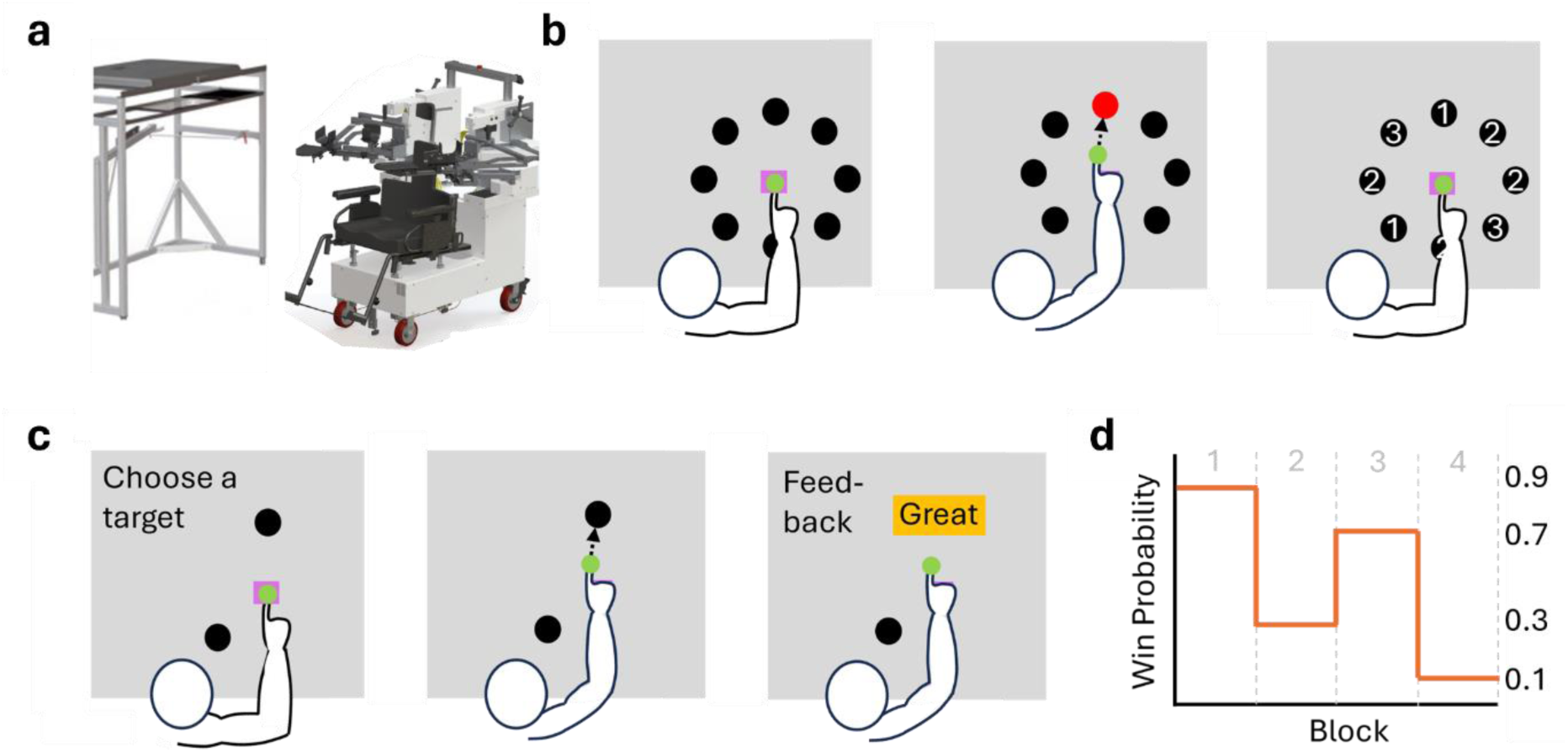
Experimental Design. **a)** Illustration of the KINARM Exoskeleton; a robot which gathers limb kinematic data during task performance. **b)** Illustration of Baseline Assessment. Participants reached from a central target to eight peripheral targets arranged in a circle (10 cm radius). They rated the difficulty of reaching each target (1–5), and the two easiest were selected for the main task. Target positions were adjusted for stroke patients to ensure reachability and prevent physical impairments from biasing choice behaviour. **c)** Illustration of Main Experiment. A reversal learning task assessed belief updating in chronic stroke patients. Participants reached for one of two preselected targets, aiming to maximise rewards while adapting to changing action-outcome contingencies. Each trial began with cursor positioning at the central target, followed by the appearance of two peripheral targets as the “go” signal. Participants chose a target and received binary feedback (“Great” or “Oh no”). Reward probabilities reversed every 18–21 trials, requiring continuous adaptation. Each limb completed 82 trials. **d)** Reward probabilities. The experiment consisted of four blocks with shifting reward probabilities. The reward probabilities for Targets 1 (T1) and 2 (T2) across Blocks 1 to 4 were as follows: 0.9/0.1, 0.3/0.7, 0.7/0.3, and 0.1/0.9, respectively. The contingency order was reversed for the other limb, and task sequence was counterbalanced across participants.

### Experimental Design and Procedures

#### 1) Baseline assessments

The baseline assessment identified accessible movement targets for each participant to ensure that subsequent choice behaviour was not confounded by motor impairment. Participants moved from a central square target (1.5×1.5 cm) positioned at mid-workspace (90° elbow flexion, 30° shoulder flexion) to eight peripheral targets (1.5×1.5 cm) evenly spaced 10 cm from the centre (**Figure 1b**). After completing all reaches three times in a row, participants rated the difficulty of each target on a scale from 1 (easiest) to 5 (most difficult). The two targets rated as the easiest were then selected for the main task. For stroke patients, adjustments were made to the positions of the central and peripheral targets as necessary to ensure they could effectively reach their selected targets **(Figure 1b)**.

#### 2) Main experiment

Participants made reaching movements toward one of the two selected peripheral targets to earn points. On each trial, one target was more likely to yield a reward, but these reward contingencies changed unpredictably over time. Participants were informed that reward probabilities were reciprocal (i.e., *P*(win | T1) = 1 – *P*(win | T2)) and would occasionally reverse. To maximise rewards, they had to integrate feedback across trials to infer which target was currently more rewarding. Importantly, there were no time constraints, and participants were not instructed to respond as quickly as possible.

Each trial began with the cursor positioned in the central target. After a brief hold (500– 550ms), the two peripheral targets appeared, serving as the go signal. Participants reached to one target (**Figure 1c**), held it for 500ms to confirm their choice, and immediately received on-screen feedback indicating success (“Great”) or failure (“Oh no”) for 800ms, followed by a 300ms inter-trial interval. Participants completed 78 trials with each upper limb.

The task structure followed that described in previous studies^32,33^. Reward contingencies reversed every 18–21 trials. For one upper limb, reward probabilities for Targets 1 (T1) and Target 2 (T2) across four blocks were 0.9/0.1, 0.3/0.7, 0.7/0.3, and 0.1/0.9 (**Figure 1d**); for the other upper, the order was reversed to avoid carry-over effects. Task order (weak vs. strong limb first) was counterbalanced across participants.

Seventeen trials into each block, participants were prompted to complete a meta-confidence probe. They were asked to indicate which target they believed was currently more rewarding and rated their confidence on a 1–5 scale (1 = very unsure, 5 = very sure). This measure served to ensure sustained engagement, verify comprehension of the task structure, and provide an index of metacognitive awareness into how participants monitored and adapted to changing reward contingencies over time.

Overall, the task was designed to provide sufficient exposure to each contingency before a reversal occurred (every 18–21 trials), allowing participants to learn the probabilistic relationships between actions and outcomes while minimising fatigue by limiting the total number of trials to 78. Simulations using the HGF confirmed that learning parameters could be robustly recovered at this task length and reversal frequency (**Supplementary Materials**, HGF details below).

The task was custom-designed in Simulink (MATLAB 2019b) and executed using KINARM Dexterit-E. Performance data were recorded at 1 kHz and included the following behavioural variables: (i) choices (T1 or T2), (ii) outcomes (rewarded or unrewarded; no timeouts were present due to unlimited response time), and (iii) metacognitive confidence ratings (Likert scale 1–5, with 5 indicating highest confidence). Post hoc behavioural analysis in MATLAB 2020b extracted key metrics for statistical analysis, allowing us to address our first research question. The metrics included (iv) the adjusted rate of correct responses, or *choice accuracy*, which accounted for the probabilistic task structure and reflected the proportion of trials in which participants selected the target with the higher reward probability on that trial, irrespective of whether a reward was actually delivered^42^, (v) the probability of staying or shifting responses after a rewarded (win) relative to an unrewarded (loss) trial, and (vi) movement time (MT, in ms). Movement time (MT) was defined as the interval from the presentation of the choice, which served as the ‘go’ signal, until the participant’s hand position landed within the chosen target.

#### 3) Control Assessment

Motor impairment was quantified using the Fugl-Meyer Upper Extremity (FM-UE) assessment, while cognitive function was evaluated with the standardised KINARM *Trail Making Task* (TMT) performed with the strong limb. The TMT (Part A) was administered to test participants’ cognitive abilities^43^ and is particularly sensitive to visual search^43^, motor speed skills^43^, processing speed^43^, and fluid cognitive ability^44^. Participants were instructed to connect 25 semi-randomly dispersed numbered targets in ascending order as ‘fast and accurately as possible’. All targets were presented simultaneously. A cursor followed participants’ hands. If participants hit an incorrect target, the target they had successfully reached before turned red (from white) and participants had to return to that target before proceeding. Before each 1-25 run, participants completed a practice run with five targets numbered 1-5. No time limits were imposed.

### The hierarchical Gaussian filter

To model individual trial-by-trial decision-making performance in our task, we used the Hierarchical Gaussian Filter (HGF), an established hierarchical Bayesian inference model^35,45^. The HGF, available as part of the open-source TAPAS software (http://www.translationalneuromodeling.org/tapas), was implemented using version 7.1 of the HGF toolbox in MATLAB 2020b (The MathWorks).

The HGF is a generative model of learning, which assumes that individuals infer hidden environmental states, such as the probabilistic mapping from stimuli (or actions) to outcomes, underlying causes of sensory inputs, and their changes over time (volatility). In our study, we used a three-level enhanced HGF (eHGF) for binary categorical outcomes^36^, to model how participants updated their beliefs about the tendency of action–outcome contingencies (level 2, latent state *x*_2_) and the volatility of those contingencies over time (level 3, log-volatility, *x*_3_, **Figure 2a**). The observed binary outcomes from the task are represented at level 1 (*x*_1_), e.g., *x*_1_*^(k)^* = 0 or 1 representing the rewarded action, T1 or T2, respectively, on trial *k*.

**Figure 2.**
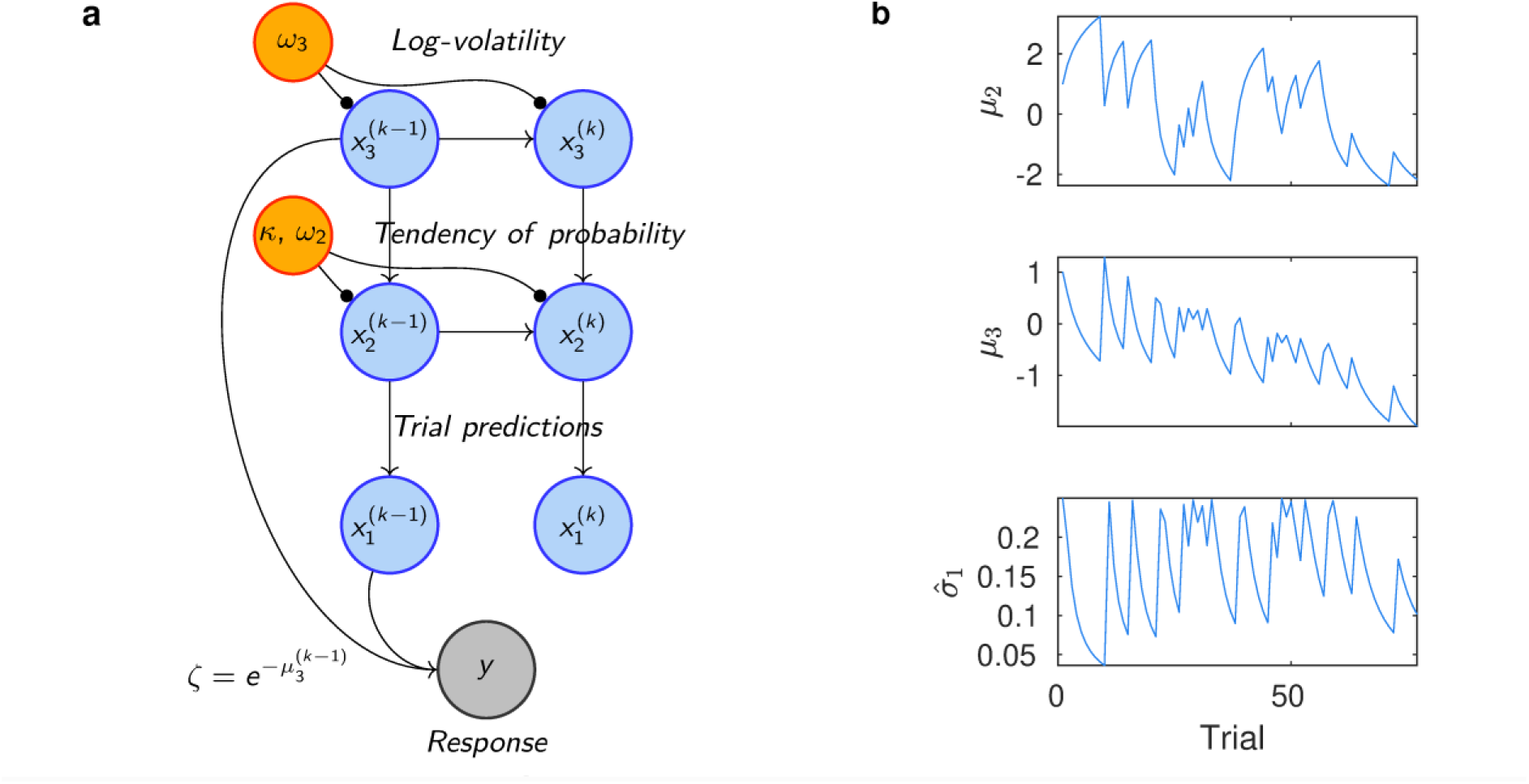
HGF computational model. **a)** Illustration of the best-fit three-level eHGF model (HGF3) with relevant parameters modulating each level (adapted from^48^). In this framework, agents infer the true state of the action–outcome contingency on trial *k, (x_2_^(k)^),* and its rate of change, or log-volatility *(x_3_^(k)^).* Beliefs about these states are modelled as Gaussian distributions, parametrised by their means (*μ_2_^(k)^*, *μ_3_^(k)^*) and variances (σ*_2_^(k)^*, σ*_3_^(k)^*), representing uncertainty (i.e., the inverse of precision). These belief parameters are updated via one-step equations, modulated by parameters such as *κ*, *ω*_2_, and *ω*_3_. The response model maps beliefs to decisions based on the expected log-volatility from the previous trial *(μ_3_^(k-1)^*), which serves as the prediction for the current trial. **b)** Illustration of trajectories of mean posterior beliefs on levels 2 (top) and 3 (center), and informational uncertainty about the outcome, σ*_1_^(k)^*, in one participant.

Beliefs at each level are defined as Gaussian distributions, described by posterior means (*μ*_2_, *μ*_3_) and variances (*σ*_2_, *σ*_3_), which represent uncertainty about the corresponding hidden states, and their inverse (π_2_, π_3_), precision of those beliefs (**Figure 2b**). At level 2, the variance *σ*_2_ is conventionally termed “informational” or “estimation uncertainty”, while *σ*_3_ is referred to as “uncertainty about volatility”. The trial-wise predicted uncertainty about outcomes, i.e., Bernoulli variance at level 1, is denoted by *σ̂*_1_.

Belief updating in the eHGF is driven by PEs, the discrepancy between expected and observed outcomes, weighted by precision ratios. These precision-weighted prediction errors (pwPEs) determine how strongly beliefs are updated in response to new evidence^35,36^. The belief update at level *2* on trial *k* is governed by the expression:

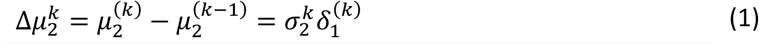

Here, the update to the tendency of the action-outcome contingencies, *μ_2_^(k)^*, is proportional to the PE about outcomes, *δ_1_^(k)^*, weighted with the estimation uncertainty on that level, *σ_2_^(k)^*, such that pwPE_2_ = *σ_2_^(k)^δ_1_^(k)^*. The full model equations, including the belief-updating equation for level 3, are provided in **Supplementary Materials**, based on the original HGF by Mathys (2011,2014)^35,36^ and Hess et al. (2025)^42^.

The evolution of hidden states *x_2_*, *x_3_* is modelled as a Gaussian random walk. In particular, log-volatility *x_3_* directly influences the time evolution of *x₂* via its conditional variance, expressed as:

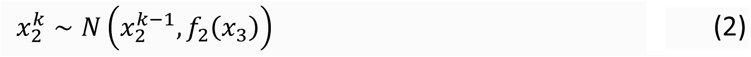

with (dropping *k* for simplicity)

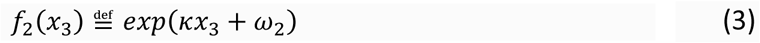

Greater *μ*_3_ corresponds to greater expectation of environmental volatility, driving faster adaptation in *μ*_2_, consistent with normative accounts of learning under uncertainty^46^.

Learning dynamics are modulated by key parameters: (i) *ω_2_*, tonic volatility at level 2, which modulates belief updating about action–outcome contingencies (smaller *ω_2_* for slower updates; **Figure S1**); (ii) *ω*_3_, tonic volatility at level 3, influencing higher-level updates in *μ_3_*; (iii) *κ*, a coupling constant that determines how phasic environmental volatility (*x*_3_) modulates learning at level 2.

To assess how beliefs mapped to decisions, we coupled this perceptual model to previously established response models^32,34,36,47^. Model M1 combined the 3-level eHGF with a unit-square sigmoid response function shaped by a fixed inverse decision noise parameter ζ, interpreted as inverse decision noise: as *ζ* increases, the sigmoid approaches a step function. Model M2 used a 2-level eHGF with constant volatility. Model M3 retained the 3-level structure but allowed the sigmoid to vary trial-by-trial based on predicted log-volatility^47^ such that higher volatility estimates produced more stochastic responses. In M1 and M3, both *ω₂* and *ω_3_* were free parameters; in M2, only *ω_2_* was free. *ζ* was estimated in M1 and M2, while in M3, initial values *μ*_3_^(0)^ and *σ_3_*^(0)^ were free, representing prior expectations about volatility and associated uncertainty. A fourth model (M4) was identical to M3 but replaced *ω*_2_ with κ^34^.

Between-group differences in these model parameters directly address our second research question. Specifically, we used *ω_2_*, ω_3_, *κ*, *μ*_3_^(0)^ and mean *μ*_3_ to assess whether stroke affects the speed of belief updating at different hierarchical levels. Smaller ω_2_ values reflect slower adaptation of action–outcome beliefs, while lower ω_3_ indicates reduced updating of log-volatility estimates. Higher initial (*μ_3_*^(0)^) and mean (*μ*_3_) log-volatility estimates indicate that individuals expect the environment to be more unstable, driving faster adaptation in *μ*_2_ (eq. 2). In M3–M4, higher trial-wise *μ_3_* values lead to a noisier mapping between beliefs and choices, reflecting greater decision stochasticity; analogous to smaller *ζ* in M1–M2. Last, larger *κ* values indicate stronger modulation of lower-level learning by volatility (eq. 3).

Models were fit to individual action–outcome series using priors in **Table S2.** Random-effects Bayesian model selection based on log model evidence was used for comparison. Parameter recoverability was assessed through simulation, following previous work^48^. All models were implemented in the TAPAS toolbox (release v7.1) using the function tapas_ehgf_binary in MATLAB R2020b.

For statistical analysis, we focused on the model parameters capturing learning dynamics (*ω_2_, ω_3_, μ_3_^(0)^*, mean *μ_3_, κ*) and decision stochasticity (*ζ* or mean *μ_3_*), depending on the winning model, consistent with previous work^32,34,38,49,50^.

### Assessing motor invigoration

To assess motor vigour, we constructed a series of Bayesian multilevel regression models with the natural logarithm of movement time (*logMT,* in log-ms) as the DV. This approach followed our previous work in clinical populations^32,34^. As in those studies, the primary continuous regressor was the *strength of predictions* about action– outcome contingencies, 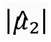. The strength of predictions is higher on trials where the absolute value of beliefs about reward probability is large, that is, when participants have a strong expectation of being rewarded for the correct action.

Although some HGF variants jointly model categorical decision-making and reaction times (RT, via linear regression; e.g.^51^), they have not been validated for group comparisons where one group responds markedly slower, as in Parkinson’s disease or stroke. Simulations using a multimodal HGF framework^51^ showed that slower RTs in a group can bias inference on decision-related parameters, leading to misidentification of faster learners as slower ones or the reverse (see **Supplementary Materials, Figure S2**). Therefore, we did not apply this complementary approach to our data and instead followed our planned analyses^32,34^, analysing MT separately using Bayesian multilevel regression models rather than incorporating it into the HGF.

In our Bayesian multilevel regressions, we built a hierarchy of models of decreasing complexity by progressively removing fixed and random effects (**Table S3**). The full model included a three-way interaction between group, limb, and the centred trial-wise predictor (*prediction.c*), with random intercepts and slopes by subject. Centring improves model stability and interpretability of the intercept. HC and the strong limb served as reference levels, so posterior estimates reflected differences relative to these baselines. Effects were considered credible when the posterior estimates and their 95% CrIs excluded zero

We also considered models that replaced the continuous regressor with trial-wise informational uncertainty at the outcome level (Bernoulli variance *σ̂*_1_), which has also been shown to explain the relationship between decision-making and movement time^51,42^. However, both 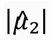 and *σ̂*_1_ were highly collinear, exhibiting a robust negative correlation of approximately –0.9 in every participant. Accordingly, we focused our main analysis on models explaining strength of prediction, and report the comparison across all models including either regressor in the **Supplementary Materials**.

Model comparison was conducted using leave-one-out cross-validation (LOO-CV) with Pareto-smoothed importance sampling^52^. The best-fitting model was identified based on the highest expected log point-wise predictive density (ELPD). We also ensured that the absolute mean difference in ELPD between the two top models (elpd_diff in brms) exceeded twice the standard error of these differences (2se_diff). If elpd_diff was smaller than 2se_diff, we opted for the more parsimonious model.

### Statistical Analysis

Dependent variables (DV: general performance indices, ΔPerceived Reaching Difficulty, metacognitive scores, computational parameters) were analysed using both frequentist and Bayesian ANOVA in a 2 × 2 factorial design with group (SK, HC) and limb (dominant (HC)/less impaired (SK) [strong], non-dominant (HC)/more impaired (SK) [weak]) as factors. We used both frequentist and Bayesian approaches to assess statistical significance and quantify the strength of evidence for or against each effect. In Bayesian ANOVAs, Bayes Factors (BF) quantified the ratio of model likelihoods between a reduced model (excluding a factor) and the full model. BF < 1 supports including the factor, whereas BF > 1 supports its exclusion, interpreted according to Wetzels and Wagenmakers (2012)^53^. Significant interactions were followed by independent-samples t-tests, reporting BF_10_ as evidence for the alternative (H₁) relative to the null (H₀). BF_10_ > 1 supports group or limb differences; BF_10_ < 1 supports their absence.

For variables with non-normal distributions (identified via Levene’s test or residual inspection), we applied non-parametric 2 × 2 factorial analyses using synchronised rearrangements^55^ (5000 permutations), followed by pairwise permutation tests (5000 permutations). P-values from non-parametric tests are provided with four decimals, as 5000 permutations yield a minimum possible value of 0.0002 (1/5000). Non-parametric effect sizes were estimated using the probability of superiority (Δ for within-subjects, Δ_sup_ for between-subjects; range 0-1)^54,55^. In frequentist analyses, *α* = 0.05 was used, and false discovery rates (FDR) were controlled at *q* = 0.05 for multiple comparisons.

To model trial-wise behaviour (stay vs shift) as a function of previous outcome (win vs loss) and other covariates, we used Bayesian logistic regression. The full model predicted the probability of staying with the same choice (stay = 1, shift = 0) from previous outcome, group, and limb, including all interactions and a random intercept for subject: stay ∼ 1 + previous_outcome * group * limb + (1 | subject). Simpler models sequentially excluded these terms, and model comparison used LOO-CV, as in the motor vigour analyses.

Standard correlation analyses used Spearman’s ρ, with results reporting ρ, *p*-values, and 95% confidence intervals (CI). Motor vigour analyses followed the Bayesian workflow framework^56^, as detailed in previous section.

## Results

### Task setup validation

The difference in perceived reaching difficulty (*ΔPerceived Reaching Difficulty*) showed a markedly skewed distribution, with Levene’s test indicating a violation of normality assumptions, necessitating a non-parametric approach. We therefore applied non-parametric synchronised rearrangements to evaluate main and interaction effects^57^.

No significant main effects of group (*p* = 0.54) or limb (*p* = 0.59), nor a group × limb interaction (*p* = 0.53), were observed. These results indicate no significant differences in perceived reachability across groups or limbs, suggesting that any observed differences in task performance are unlikely to be confounded by perceived reaching difficulty.

### Reduced reward-based learning in stroke participants

Factorial analysis of the choice accuracy revealed a significant main effect of group, with SK showing lower choice accuracy than HC, indicating an impaired ability to choose the most likely rewarding option over time, (*F*(1, 136) = 12.32, *p* = 0.0006, η² = 0.081, BF = 0.02; **Figure 3a**). There was no significant main effect of limb (*F*(1, 136) = 2.05, *p* = 0.15, η² = 0.014, BF = 5.21), nor a significant group × limb interaction (*F*(1, 136) = 1.18, *p* = 0.28, η² = 0.0077, BF = 2.8).

**Figure 3.**
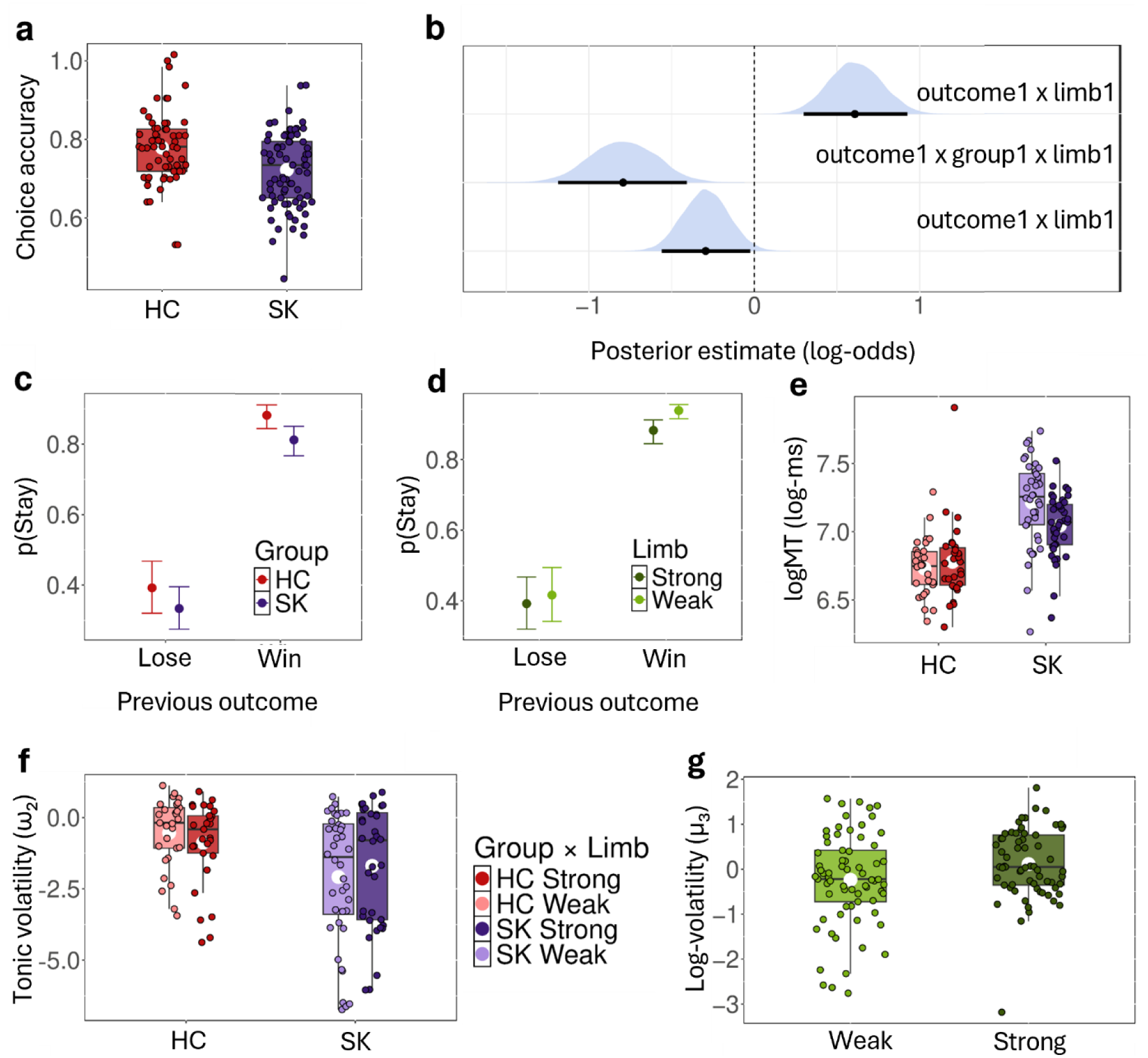
Reward-based learning impairment shows specificity to the weak limb in stroke. Boxplots **a** and **e-g** show individual and group-level data for stroke participants (SK, purple) and healthy controls (HC, red), separated by strong (darker shades) and weak (lighter shades) limbs. White dots indicate group means. **a)** SK showed a significantly lower *choice accuracy* than HC, with no significant main effect of limb or interaction (2×2 limb × group ANOVA). **b)** Posterior distributions (blue) and 95% credible intervals (CrIs) for selected fixed effects of the Bayesian logistic regression assessing the probability of staying as a function of previous outcome. Displayed effects (and others reported in the main text) were considered credible, as their 95% CrIs did not include zero. Notation: “outcome1” contrasts win minus loss, on the previous trial; “group1” denotes SK–HC; “limb1” contrasts weak minus strong limb. The negative posterior estimate for outcome1 x group1 x limb1 provides evidence for a three-way interaction, indicating that the effect of previous outcome on staying probability depends jointly on group and limb. **c)** Estimated marginal probabilities of staying as a function of previous outcome (win/lose) and group, showing the posterior mean (dot) and 95% CrI. **d)** Same as panel c, but for the interaction of previous outcome with limb1. **e)** A credible group × limb interaction was observed for log-transformed movement time (*logMT*, in log-ms), driven by slower performance with the weak limb in SK. This effect was absent in HC. SK also showed significantly slower movement times overall (group effect), with strong support from both frequentist and Bayesian analyses. **f)** *Tonic volatility (ω_2_)* showed a significant group × limb interaction, with SK participants displaying lower *ω_2_* in their weak limb compared to their strong limb—a pattern not observed in HC. Additionally, *ω*_2_ was lower in SK than HC for the weak limb only, suggesting reduced tonic learning rates and more rigid belief updating in stroke. **g)** Mean *log volatility estimates (μ_3_)* were lower for the weak limb across both groups, indicating an underestimation of environmental volatility. No group or interaction effects were observed.

To assess whether stroke affected the use of reward feedback in guiding decisions, we modelled the probability of staying with the same choice as a function of previous outcome (win/loss), group, and limb. The best-fitting Bayesian logistic regression included all interactions among these factors and a random intercept for subject (LOO-CV: elpd_diff = 24.07 vs. next-best model, exceeding 2 × se_diff = 14.53). Posterior estimates from this model are shown in **Figure 3b–d**.

To aid interpretation, we note that the following reference levels were used in the model: outcome1 contrasts win (1) versus loss (0), with loss as the reference; group1 contrasts stroke (1) versus control (0), with healthy controls as the reference; and limb1 contrasts the weak (1) versus strong (0) limb, with the strong limb as the reference. In this context, the effect of outcome1 reflects how much more likely participants are to stay after a win compared to a loss, the effect of group1 captures overall differences in behaviour between stroke survivors and healthy controls, and the effect of limb1 reflects differences in behaviour when using the weak versus the strong limb.

In the reference condition (HC using strong limb), there was a robust win–stay tendency relative to loss–stay (log-odds for outcome1 = 2.46, 95% CrI [2.25, 2.67]). The intercept for stay after a loss was –0.44 (probability = exp(–0.44)/(1 + exp(–0.44)) = 0.39), while the estimated probability of staying after a win increased to ∼0.88 (log-odds = –0.44 + 2.46 = 2.02; probability = exp(2.02)/(1 + exp(2.02))). This win–stay effect was credibly reduced in stroke participants (outcome1 x group1 = –0.29, 95% CrI [–0.56, –0.03]) and further attenuated when using the weak limb (three-way interaction: outcome1 x group1 x limb1 = –0.79, 95% CrI [– 1.19, –0.41]), indicating a specific disruption in reward-guided decision-making in the impaired effector. By contrast, healthy controls showed an enhanced win–stay effect when using the weak (non-dominant) limb (outcome1 x limb1 = 0.61, 95% CrI [0.30, 0.93]).

In summary, general decision-making performance showed robust differences between groups in accuracy rate and *win–stay* tendencies, with additional credible interactions in the probability of staying after a win, driven by SK participants being more likely to shift responses after receiving a reward especially when using their weak limb.

### Meta-Confidence Does Not Differ Between Stroke and Control Groups

To assess whether group differences in reward-based learning were attributable to differences in metacognitive awareness, we analysed average *meta-confidence* ratings across blocks using a mixed ANOVA. There were no significant main effects of group (F(1, 68) = 0.990, p = 0.323, partial η² = 0.063; BF = 5.05) or limb (F(1, 68) = 2.34, p = 0.130, partial η² = 0.033; BF = 2.56), and no significant group × limb interaction (F(1, 68) = 0.012, p = 0.913, partial η² < 0.001; BF = 8.33). These results suggest that meta-confidence was comparable across groups and limbs, with moderate to strong Bayesian evidence favouring a model excluding those factors.

### Motor and cognitive function do not explain impaired reward-based learning after stroke

Furthermore, in control analyses, we examined whether general performance during reward-based learning in stroke survivors related to motor or cognitive impairment. We correlated the adjusted rate of correct responses with two clinical measures: the *FM-UE* (motor function) and the *TMT* from the KINARM battery (cognitive function). Associations with motor scores were assessed for the weak limb only, as *FM-UE* data were not available for the strong limb; conversely, *TMT* performance was analysed for the strong limb, since the task is administered with that arm to minimise motor confounds.

Correlation analyses revealed no significant associations between motor function (*FM-UE* scores) and adjusted correct response rate (Spearman’s ρ = 0.064, *p* = 0.693, 95% CI [–0.231, 0.371]; BF = 0.40). For cognitive function, there was also no significant association, though anecdotal BF evidence suggested a weak trend: lower (better) TMT scores correlated with higher adjusted correct response rate (ρ = –0.221, *p* = 0.209, 95% CI [–0.507, 0.106]; BF = 1.91).

We further assessed whether motor function in stroke explained changes in the probability of staying after a win relative to a loss when using the weak arm. Bayesian logistic regression revealed no credible effect of FM-UE scores on the reference probability of staying after a loss (posterior slope = 0.012, 95% CrI [−0.031 0.056], log-odds scale). Likewise, there was no credible interaction effect between motor function and previous outcome, as the regression coefficient for the difference in slopes between win and loss trials overlapped with zero (outcome1 x motorfunction posterior slope = −0.012, 95% CrI [−0.030 0.007]), indicating that the extent of motor impairment did not modulate the influence of reward on stay behaviour.

By contrast, TMT scores showed a credible negative effect on the probability of staying after a loss with the strong limb (posterior estimate = –0.389, 95% CrI [–0.725, –0.052]), indicating that poorer cognitive performance (higher TMT scores) was associated with a reduced tendency to stay after a loss. Although this effect was unexpected, there was no credible interaction between TMT scores and previous outcome (outcome1: x cognitiveperformance slope = 0.031, 95% CrI [–0.217, 0.280]). Thus, cognitive performance did not explain differences in stay behaviour between wins and losses. Instead, participants with poorer cognitive performance were less likely to repeat the same response, regardless of the previous outcome. This pattern was confirmed using a Bayesian logistic regression assessing *p*(stay|win) in the strong limb as a function of TMT scores (slope = –0.343, 95% CrI [–0.644, – 0.045], log-odds scale).

Together, these findings indicate limited evidence that general motor or cognitive function accounts for variability in overall reward-based learning performance among stroke survivors. Although poorer cognitive performance in the strong limb was associated with a decreased tendency to repeat responses—reflecting reduced behavioural “stickiness”—this effect did not depend on previous outcomes.

### Stroke patients move slower particularly with their weak limb

Looking at general motor performance in this task, we analysed *log-transformed MT* (log-ms) to examine group and limb effects (**Figure 3e**). A 2 x 2 factorial analysis revealed a significant group × limb interaction (*F*(1, 136) = 5.91, *p* = 0.016, partial η² = 0.028, BF = 0.30). Post-hoc tests showed SK participants were slower with their weak limb (*p* = 0.007, *d* = 0.624, BF = 6.23), while HC showed no difference between limbs (*p* = 0.45, BF = 0.33). There was overwhelming evidence for SK participants showing overall slower *log-transformed MT* than HC (group effect, *F*(1, 136) = 69.84, *p* < 0.0001, partial η² = 0.325, BF ≈ 0). No main effect of limb was found (*p* = 0.090, BF = 2.33). Together, as expected, these results indicate clear group differences in movement time, with SK participants showing greater slowing, particularly when using their weak limb.

### Reduced reward-based learning in stroke reflects slower belief updating about action-outcome contingencies in the weak limb, not differences in estimated environmental volatility

To investigate whether stroke survivors exhibit altered decision-making under uncertainty, we analysed key computational variables from the winning model identified by Bayesian Model Selection (BMS; see *Methods*). BMS revealed that model M3 best explained participants’ behaviour in the joint sample for both weak and strong limbs (exceedance probability, *P_exc_* = 1; expected frequency, *F_e_*_xp_ = 0.762 and 0.707, respectively), as well as within each group independently (HC, weak limb: *P_exc_* = 0.995, *F_e_*_xp_ = 0.738; strong limb: *P_exc_* = 0.970 *F_e_*_xp_ = 0.592; SK, weak limb: *P_exc_* = 0.997, *F_e_*_xp_ = 0.690; strong limb: *P_exc_* = 1.000, *F_e_*_xp_ = 0.725).

In this model, a higher expectation of *log-volatility* on a given trial is associated with increased decision noise (lower inverse decision temperature) leading to a more stochastic mapping from beliefs to choices. The BMS results indicate that, among the tested models, behaviour was best captured by a hierarchical model in which expected environmental volatility dynamically modulates the belief-to-decision mapping. Simulation analyses confirmed good parameter recovery (**Figure S3**).

Factorial analyses of *tonic volatility* on level 2 (*ω_2_*), level 3 (*ω_3_*), initial (*μ_3_^(0)^*) and mean phasic log-volatility estimates (*μ_3_*) were conducted using non-parametric synchronised rearrangements^57^, as these variables exhibited skewed distributions (confirmed via Levene’s test). A significant main effect of group was observed for *ω_2_* (*p* = 0.0120), along with a significant group-by-limb interaction (*p* = 0.0086; **Figure 3f**). Post hoc analyses showed that SK participants exhibited significantly lower *ω_2_* in their weak limb compared to their strong limb (permutation test, *p*_FDR_ = 0.0130, *Δ* = 0.7000, large effect size), an effect not observed in HC (p = 0.2020). Additionally, *ω_2_* was significantly lower in SK than in HC for the weak limb (*p*_FDR_ = 0.0014, Δ_sup_ = 0.7242), but no group differences were found for the strong limb (*p* = 0.15). Since lower *ω*_2_ reflects reduced tonic learning rates (i.e., smaller updates in *μ*_2_), these results suggest more rigid updating of probabilistic contingencies in the weak limb of stroke survivors. No main effect of limb was observed (*p* = 0.4146).

In addition, a significant main effect of limb was observed for *μ_3_* (*p* = 0.0076), with participants underestimating volatility in the weak limb relative to the strong limb (p = 0.0034, Δ = 0.6286). This effect was independent of group, as no significant group effect was found (*p* = 0.09), nor was there evidence of a group-by-limb interaction (*p* = 0.2624; see **Figure 3g**). Last, no significant main effects of group, limb or their interaction were observed for *μ_3_ ^(0)^* or *ω_3_* (*p* > 0.10 in all cases).

To determine whether these *ω_2_* findings could be attributed to general impairment, we examined correlations between *ω_2_* and both overall motor function (*FM-UE)* and cognitive performance (*TMT)*. No association was found between *ω_2_*and either motor function (FM-UE: ρ = –0.147, *p* = 0.366, 95% CI [–0.441, 0.161], BF = 0.56) or cognitive performance (TMT: ρ = –0.202, *p* = 0.253, 95% CI [–0.522, 0.161], BF = 0.79), indicating that reduced tonic volatility in stroke reflects specific alterations in learning dynamics rather than general motor or cognitive impairment.

### Strength of predictions about reward contingencies invigorates movement similarly in stroke and control participants

We examined whether trial-wise strength of predictions about action-outcome contingencies (**Figure 4a**) invigorate movement in stroke survivors and healthy controls, and whether the sensitivity of this association decreased in SK relative to HC groups. Model 9 emerged as the best-fitting model, outperforming the next-best alternative (model 8; elpd_diff = 182.88, > 2 x se_diff = 2 x 19.16; substantial improvement). Posterior estimates for model effects are presented in **Table 2**. Posterior predictive checks confirmed that the observed logMT values overlapped well with simulated datasets from the posterior distribution (**Figure S4**).

**Figure 4.**
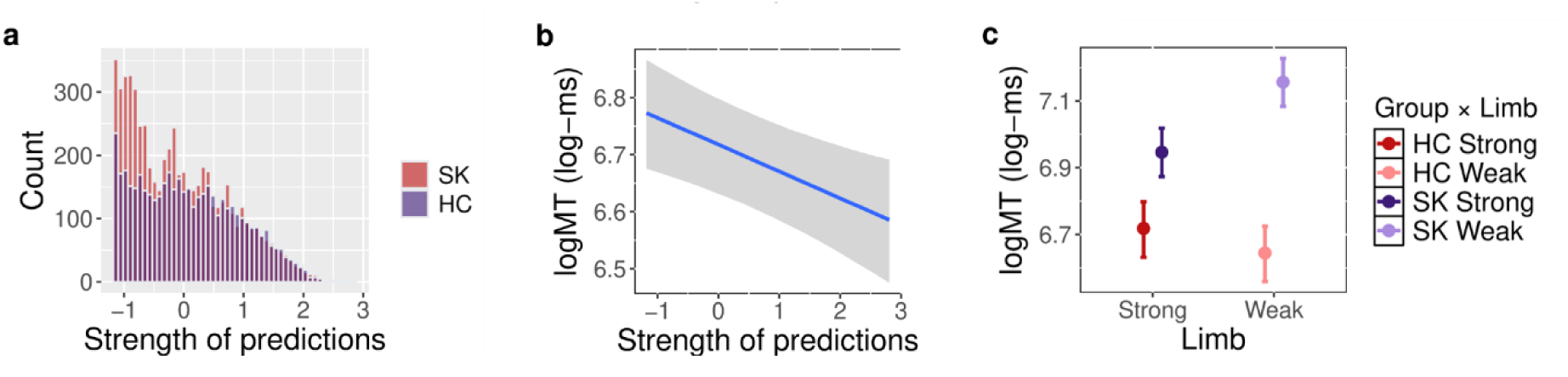
Invigoration of movement time is associated with strength of predictions about the tendency of the action-reward contingency across SK and HC groups. **a**) Distribution of the centred regressor used in hierarchical Bayesian regression modelling: trial-wise strength of predictions about the action-reward contingency, 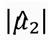, in SK (purple) and HC (red) groups. **b**) In the best-fit model, model 9 (see Table 2 for a complete report of posterior estimates), there was a negative association between 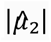, and log-transformed movement time (*logMT*, in log-ms), indicating faster movements under strong predictions about the reward contingency. Posterior estimate for linear regression with 95% credible interval (CrI). **c**) group × limb credible interaction on baseline *logMT*, indicating relatively slower performance with the weak than strong limb in stroke participants compared to HC, as expected from Figure 3e. Posterior estimates and 95% CrI for logMT (log-ms) for each group and limb are indicated by circles and error bars.

**Table 2.**
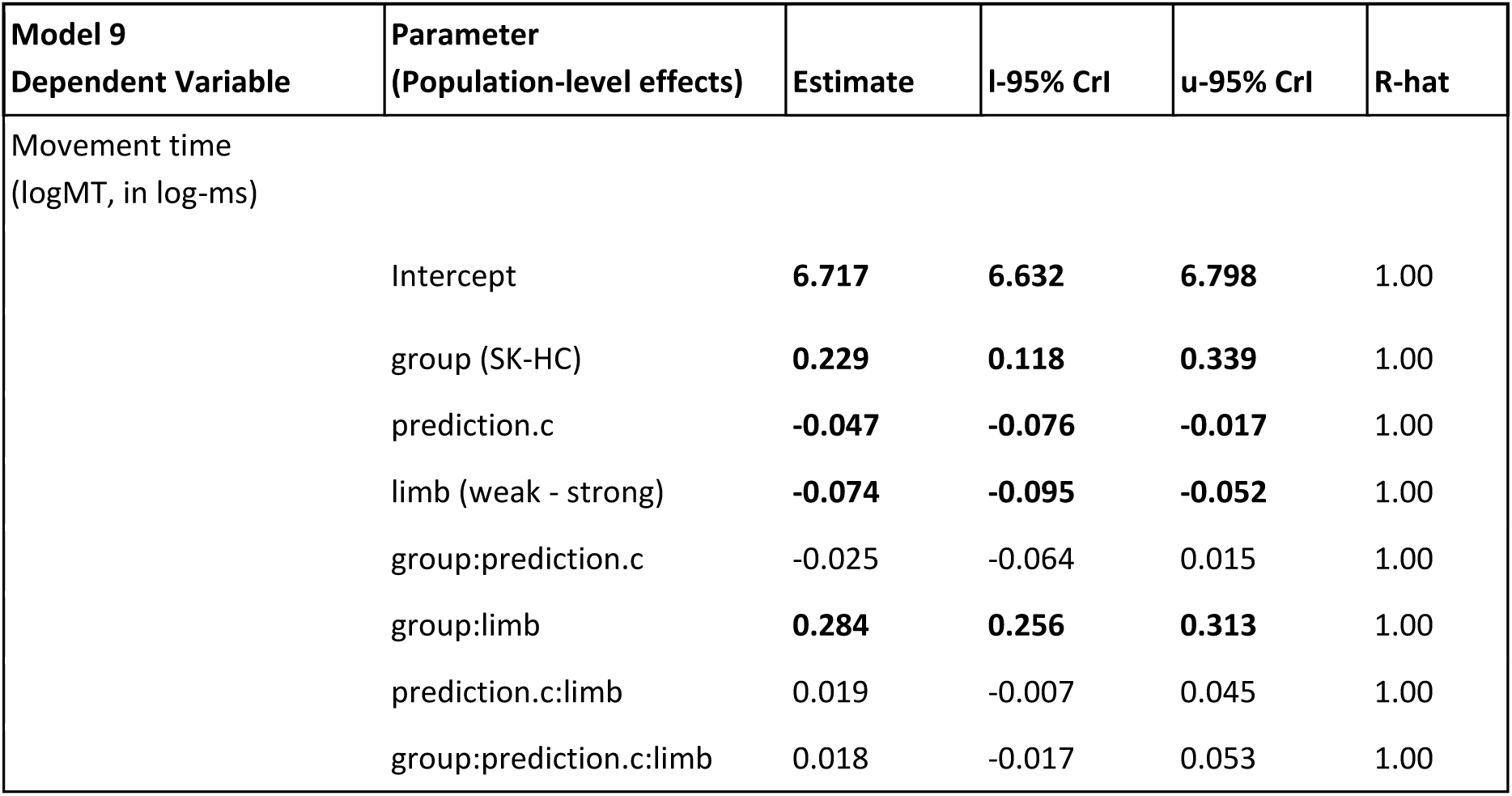
Summary parameter estimates for the winning Bayesian multilevel model assessing the effect of strength of predictions about the tendency of the action-reward contingency on timing performance. Estimate = posterior mean; CrI = credible interval based on quantiles. Gelman-Rubin statistics demonstrate excellent chain convergence (R-hat < 1.01). The effective sample size (ESS) was >> 400 for each parameter estimate, denoting good convergence. The predictor “prediction.c” denotes the centred values of the absolute value of beliefs about action-outcome contingencies, 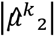. Credible effects are considered when the 95% lower and upper-bound CrI around the Bayesian point estimate do not include zero (denoted by bold font).

| Model 9<br>Dependent Variable | Parameter<br>(Population-level effects) | Estimate | l-95% CrI | u-95% CrI | R-hat |
| --- | --- | --- | --- | --- | --- |
| Movement time<br>(logMT, in log-ms) |  |  |  |  |  |
|  | Intercept | **6.717** | **6.632** | **6.798** | 1.00 |
|  | group (SK-HC) | **0.229** | **0.118** | **0.339** | 1.00 |
|  | prediction.c | **-0.047** | **-0.076** | **-0.017** | 1.00 |
|  | limb (weak - strong) | **-0.074** | **-0.095** | **-0.052** | 1.00 |
|  | group:prediction.c | -0.025 | -0.064 | 0.015 | 1.00 |
|  | group:limb | **0.284** | **0.256** | **0.313** | 1.00 |
|  | prediction.c:limb | 0.019 | -0.007 | 0.045 | 1.00 |
|  | group:prediction.c:limb | 0.018 | -0.017 | 0.053 | 1.00 |

Results showed that greater trial-wise strength of predictions was associated with faster movement times, indicated by a negative coefficient for *’prediction.c’* (posterior estimate = −0.047, 95% CrI [−0.076, −0.017]), with the 95% CrI excluding zero (**Figure 4b**). This effect reflects the association in the reference group (HC) and limb (strong). Group factor also had credible main effects on *logMT* (0.229, 95% CrI [0.118, 0.339]), indicating generally slower movements. In HC participants, the weak limb was associated with slightly faster movements than the strong limb (–0.074, 95% CrI [–0.095, –0.052]). Among interactions, only the group × limb effect was credible, with SK participants showing slower movements with the weak limb relative to the strong limb, compared to HC (0.284, 95% CrI [0.256, 0.313]; **Figure 4c**). These effects were expected based on our standard behavioural analysis shown in previous sections (**Figure 3e**).

By contrast, the association between predictions and MT was not credibly modulated by group, limb, or their interaction, as all posterior estimates and 95% CrI overlapped with zero. No other credible effects were observed.

Additional models including estimates of informational uncertainty about reward outcomes (*σ̂*_1_; *uncertainty.c*) as the continuous regressor performed equivalently to those using *prediction.c* (based on LOO-CV), as expected given their high collinearity. Model 9 with *uncertainty.c* yielded comparable fit to its *prediction.c* counterpart (elpd_diff = 0.79, below 2 × se_diff = 2 × 2.79) and revealed a credible effect of greater trial-wise uncertainty being associated with slower movement times (**Table S4**), with no credible modulation by group or limb.

Together, these results indicate that strong predictions about the tendency of the action-outcome contingencies invigorate movements on a trial-by-trial basis, and support that this effect is preserved in stroke. Moreover, the findings suggest that group differences in motor performance are better accounted for by baseline slowing and limb-specific impairments, rather than by altered sensitivity to strength of predictions about reward contingencies.

## Discussion

In a volatile probabilistic learning task requiring reaching movements to express choices, stroke survivors showed reduced reward-based learning compared to healthy controls, with lower choice accuracy and a greater tendency to switch after wins—particularly with the weak limb^32,34^. Healthy participants maintained successful strategies such as repeating rewarded actions, whereas stroke survivors were more likely to switch after success, suggesting a breakdown in the integration of reward outcomes that normally stabilise future choices. Importantly, this deficit was not explained by general motor or cognitive deficits: neither motor scores (FM-UE) nor cognitive measures (TMT-II) accounted for the outcome-specific effects.

Computational modelling provided a complementary account of these behavioural effects. Behaviour across groups was best captured by a hierarchical Bayesian inference model in which expected environmental volatility dynamically modulated the mapping from beliefs to choices. Within this framework, stroke survivors showed reduced tonic volatility estimates (*ω*_2_) for the weak limb relative to both their strong limb and controls. Lower *ω*_2_ indicates slower belief updating about the tendency of the action-reward association, consistent with more rigid integration of probabilistic outcomes. By contrast, higher-level volatility parameters (μ_3_, ω_3_) were comparable between groups. These findings suggest that impaired reward-based learning in stroke patients in this study is explained by changes in belief updating about action–reward contingencies, with no evident disruption to beliefs about environmental volatility.

Despite impairments in decision-making, reward-based invigoration was preserved in chronic stroke, with no effects of group or limb. Defined here as the tendency to move faster when holding strong trial-by-trial expectations about reward probability^32,34^, this form of motivational drive remained intact. These findings reveal a behavioural dissociation of reward-dependent effects in stroke: while reward expectations continue to energise movement, reward outcomes fail to be properly integrated during decision-making.

### Reconciling mixed findings in the literature

This dissociation helps reconcile inconsistencies in the in the literature on reward processing after stroke. Studies using the Monetary Incentive Dela task in the acute phase report blunted sensitivity to small, fixed rewards^27,28^, which tends to normalise within months after stroke^29^. In our paradigm, movement invigoration driven by the strength of beliefs about reward probability was preserved in chronic stroke participants across both limbs. These findings suggest that reward-based invigoration may rebound over time, although this trajectory remains to be formally characterised in longitudinal studies.

By contrast, reward-based learning in a volatile, uncertain environment remains impaired well into the chronic stage. Consistent with Lam (2016) and Sánchez-Kuhn (2025) we found that chronic stroke survivors were less likely than controls to exploit rewarded actions in probabilistic classification or reversal tasks^24,25^. Notably, preserved metacognitive awareness in our cohort suggests that while explicit inference about action–reward contingencies was partly intact, it was insufficient to support the flexible, trial-by-trial updating needed to stabilise behaviour; a result consistent with the observed reduction in *ω*_2_.

Apparent inconsistencies with research reporting preserved reinforcement-based adaptation in chronic stroke^26^ may be attributed to task design. In stable environments where reinforcement is predictable, the speed of belief updating (e.g. via *ω_2_*) may not hinder performance, as consistent contingencies reduce the need for flexible inference. In volatile contexts such as the present task, however, the ability to rapidly update beliefs is essential. Thus, the impact of stroke on reward-based learning may critically depend on the computational demands of the environment.

### A dissociation between vigour and learning

Our results suggest that stroke can lead to dissociable effects on reward processing: preserved reward-based invigoration alongside impaired probabilistic reward–outcome learning. A similar dissociation has been demonstrated in healthy participants under pharmacological manipulation, where D_2_ receptor blockade selectively reduced movement vigour without impairing reward-based learning in a sequence-learning task^11^. Likewise, Manohar et al. (2017) showed that both contingent (performance-dependent) and non-contingent (random) rewards increased response vigour, but only contingent rewards enhanced accuracy and learning^58^.

Comparable dissociations have also been reported in psychiatric populations. Patients with schizophrenia exhibit preserved hedonic experience but attenuated win–stay behaviour, reflecting abnormal weighting of reward prediction errors^50,59,60^. Similarly, in euthymic bipolar disorder, patients exhibit intact reward-driven invigoration but reduced win–stay behaviour and slower belief updating during decision making^34^, explained by alterations in HGF parameters.

In stroke, Sánchez-Kuhn et al. (2025) similarly reported reduced win–stay behaviour associated with frontostriatal lesions, and therefore, damage to these circuits may attenuate belief updating by reducing ω_2_^25^. While our findings align with this interpretation, lesion data in the present cohort were limited to broad anatomical descriptions, preventing direct structure–function inferences. Future computational neuroimaging work should formally test this hypothesis by linking lesion location and connectivity patterns to individual learning parameters within hierarchical Bayesian models.

### Dynamic motor invigoration remains intact after stroke

Movements can be conceptualised as economic trade-offs in which vigour reflects the effort the brain is willing to invest to obtain reward, typically expressed as increased movement speed, amplitude, or force^12,61^. In our study, despite overall motor slowing, stroke survivors modulated their movement speed according to their dynamic beliefs about reward probability. Across participants, strong trial-by-trial predictions about the action–reward contingency were associated with faster movement times. This effect replicates recent findings in healthy and clinical populations^32,34^ and confirms that motor vigour scales with inferred reward probability. Bayesian multilevel regression further showed that the sensitivity of this association was comparable across groups and limbs, indicating that the reward-driven invigoration of movement is preserved in chronic stroke despite slower overall performance.

A complementary effect was observed for uncertainty: greater outcome uncertainty predicted slower movements, similarly across groups. Extending earlier studies that examined deterministic or fixed reward magnitudes^10,12,13,62^, these findings demonstrate that under volatile conditions individuals use inferred expectation on reward probability—and its associated uncertainty—to dynamically regulate movement vigour. They also align with computational models linking belief updating and outcome uncertainty in hierarchical Bayesian inference to RT during decision-making^23,42,51^, and with delta-rule (Rescorla– Wagner) frameworks relating reward-related computations to peak reaching velocity^63^.

The preservation of this process suggests that motivational drive, and its putative neural underpinnings, remain largely functional in chronic stroke. Dopaminergic activity within basal ganglia circuits has been proposed to mediate motor vigour responses^61,64,65^, as shown in animal physiology^66^ and human studies^20,67^, and dopamine-replacement therapy restores vigour in Parkinson’s disease^32,68^, paralleling pharmacological manipulations in healthy participants^11,69,70^. Together, these findings indicate that while stroke disrupts reward-based learning, the motivational system translating reward expectation into movement vigour— putatively mediated by dopaminergic transmission—may not be compromised.

### Reduced reward-based learning reflects slower belief updating about action-outcome associations, not about their volatility

We characterised decision-making behaviour using a validated hierarchical Bayesian inference framework, the HGF, which generates dynamic and adaptive learning rates to capture how agents learn under uncertainty in volatile environments^35,36^. The HGF is grounded in influential theories of cortical function proposing that the brain continuously makes and refines predictions about the world through approximate Bayesian inference^37,71^. Within this framework, posterior beliefs that generate predictions are updated through PEs weighted by a precision ratio, where precision corresponds to the inverse of uncertainty in the belief distribution.

Within the HGF framework^35,36^, tonic volatility *(ω_2_)* governs the rate at which agents adjust beliefs about action–outcome contingencies in response to new evidence. The reduced ω_2_ observed for the weak limb in stroke survivors therefore reflects slower belief updating, consistent with more rigid learning dynamics. By contrast, higher-level volatility estimates *(μ_3_,)* and tonic volatility governing their updates *(ω_3_)* were comparable between groups, indicating that this deficit does not reflect altered estimates of environmental instability but rather a selective impairment in how new outcomes shape ongoing expectations about the probability of reward.

Neuroscientific evidence supports the HGF framework, identifying neural markers of PEs and precision encoding, and linking these computations to neuromodulatory systems. In particular, dopaminergic and noradrenergic signalling have been implicated in belief updating about stimulus–outcome contingencies and in the processing of uncertainty that drives such updating^21,23,51^. Cortical regions including the prefrontal cortex, anterior cingulate cortex, and orbitofrontal cortex are also involved in these computations^46,72,73^, and dysregulation within these networks has been observed across psychiatric conditions^34,74^. A key hypothesis for future work is that altered dopamine–noradrenaline balance, together with disrupted ACC– OFC–PFC interactions, may underlie the observed impairments in reward-guided decision-making in stroke survivors.

### Motor and cognitive factors do not account for reduced reward-based learning

Perceived reaching difficulty did not differ between limbs and groups highlighting that motor impairment did not bias choice. Additionally, reward contingencies reversed across blocks, eliminating the possibility of persistent movement bias. In addition, FM-UE scores were not associated with win–stay behaviour or the key computational variables of learning. Although poorer TMT performance was linked to a general tendency to switch more often, this effect was independent of reward outcome and TMT was not associated with tonic volatility *ω_2_*.

Nevertheless, cognitive load may still modulate the weak-limb effect. Controlling an impaired limb imposes greater attentional and monitoring demands, as stroke survivors often compensate by recruiting additional degrees of freedom to achieve similar movements^75^. This increased control complexity requires continuous online correction, which in healthy individuals taxes the working-memory and executive resources that normally support motor learning and adaptation^76,77^. Converging evidence from traumatic brain injury also shows that higher cognitive workload during sustained attention tasks diminishes performance^78^.

Future studies should test this hypothesis directly by systematically manipulating cognitive load during reward-based learning tasks.

### Linking disrupted reward-based learning to non-use and reduced confidence of the weak limb in chronic stroke

Our finding that stroke survivors fail to integrate rewarded outcomes properly, particularly with the weak limb, may contribute to the well-described phenomenon of “non-use”^79^. In chronic stroke, patients’ self-assessments align well with their actual performance for the strong limb, but this correspondence breaks down for the weak limb, where patients underestimate their abilities^80^. This mismatch may arise because successful outcomes with the weak limb are not effectively integrated into updated beliefs about its utility, leading to persistently low confidence and reinforced reliance on the strong limb. Thus, impaired reward-based belief updating may contribute to non-use by weakening confidence in the weak limb; a hypothesis that should be tested directly in future work.

Supporting this view, recent bimanual choice studies show that stroke survivors tend to prefer the strong limb even when the weak limb would be more rewarding^81^. This bias emerges mainly under speeded conditions, suggesting that strong-limb reliance becomes habitual when deliberate evaluation is limited. Together, these findings point to a cycle in which impaired learning of action-reward associations prevents successful weak-limb outcomes from building confidence, allowing strong-limb habits to dominate behaviour.

### Clinical and Technological Implications

Our results suggest that reward and motivational feedback can be harnessed to boost engagement, compliance, and therapy intensity which are key drivers of rehabilitation success^30^. Yet, because our stroke survivor sample showed reduced reward-based learning, therapy should combine graded, performance-contingent feedback (e.g., points or scores delivered immediately^58,82,83^) with progressive shaping schedules that gradually increase reward density^49,84^.

These principles have direct relevance for virtual reality and technology-enhanced rehabilitation, where gamification, adaptive difficulty, and reward systems are already integral. Our findings provide a mechanistic rationale for these tools: exploit intact motivational drive to sustain effort and deliver adaptive feedback to scaffold impaired reward learning thereby rebuilding confidence and counteracting non-use.

### Limitations

Stroke is a heterogeneous condition, and our results represent group-level averages rather than uniform individual profiles. The magnitude and nature of reward-learning impairments likely depend on lesion site and network disruption. Although referral letters provided broad anatomical information, we did not have access to neuroimaging data to independently verify lesion sites. Future work combining computational modelling with structural and functional imaging will be essential to link specific disruptions in belief updating (e.g., reduced ω_2_) to damage within defined frontostriatal and cortical–cerebellar circuits.

## Acknowledgements

This work was supported by the Jon Moulton Charity Trust and the BBSRC (grant reference number: 575197)

## Data availability

The data that support the findings of this study are available on request from the corresponding author. The data are not publicly available due to them containing information that could compromise the privacy of research participants.

## Code availability

Code for the present study is available at OSF upon publication.

