## Supplementary Material for "Impaired Reward-Based Learning but Preserved Motor Invigoration in Chronic Stroke"

| Patient Number | Gender | Pre-stroke Handedness | Affected upper limb | Age (years) | Time pre-stroke (months) | FMA-UL (total 54 points) | SIS | Lesion |
|---|---|---|---|---|---|---|---|---|
| 1 | M | Right | Left | 76-80 | 19 | 34 | 17 | Right parietal intracranial haemorrhage |
| 2 | M | Left | Left | 56-60 | 24 | 43 | 10 | Right hemisphere stroke (unspecified) |
| 3 | M | Right | Right | 56-60 | 39 | 33 | 20 | Left hemisphere stroke (unspecified) |
| 4 | F | Right | Right | 56-60 | 19 | 31 | 23 | Left intracranial haemorrhage |
| 5 | M | Right | Right | 26-30 | 49 | 18 | No data | Left intracranial haemorrhage |
| 6 | M | Right | Right | 51-55 | 25 | 30 | 19 | Right hemisphere stroke (unspecified) |
| 7 | M | Left | Right | 51-55 | 15 | 44 | 8 | Right infarct in the distal left internal carotid artery (ICA) and left middle cerebral artery (MCA) territory |
| 8 | M | Left | Left | 56-60 | 19 | 12 | 20 | Right hemisphere stroke (unspecified) |
| 9 | M | Right | Left | 66-70 | 27 | 22 | 9 | Right lentiform nucleus infarct |
| 10 | M | Right | Left | 56-60 | 13 | 26 | 10 | Right frontal lobe infarct |
| 11 | F | Right | Right | 16-20 | 19 | 21 | 10 | Left striatocapsular infarct |
| 12 | M | Right | Left | 36-40 | 11 | 16 | 5 | Right MCA territory stroke |
| 13 | F | Right | Right | 36-40 | 36 | 32 | 15 | Left MCA territory stroke |
| 14 | M | Right | Right | 56-60 | 41 | 46 | 13 | Left hemisphere haemorrhagic stroke |
| 15 | M | Right | Right | 71-75 | 36 | 36 | 7 | Left MCA territory stroke; lacunar infarct of posterior internal capsule; additional lacunar infarct in left thalamus |
| 16 | M | Right | Left | 46-50 | 27 | 28 | 12 | Right total anterior circulation stroke |
| 17 | M | Right | Right | 36-40 | 15 | 41 | 19 | Left internal capsule infarct |
| 18 | M | Left | Left | 71-75 | 28 | 24 | 13 | Right striatocapsular infarct |
| 19 | F | Left | Left | 61-65 | 26 | 38 | 15 | Right hemisphere stroke (unspecified) |
| 20 | M | Right | Right | 36-40 | 15 | 28 | No data | Left basal ganglia intracerebral haemorrhagic stroke |
| 21 | M | Right | Left | 21-25 | 8 | 35 | 7 | Right basal ganglia stroke |
| 22 | F | Left | Left | 51-55 | 30 | 15 | 9 | Right MCA territory stroke |
| 23 | M | Right | Left | 21-25 | 126 | 23 | 10 | Right frontal intracerebral infarct |
| 24 | M | Right | Left | 36-40 | 23 | 32 | 15 | Right MCA territory stroke |
| 25 | M | Right | Right | 26-30 | 17 | 33 | 10 | Left anterior limb of internal capsule infarct |
| 26 | F | Right | Right | 56-60 | 65 | 23 | 16 | Left lacunar infarct |
| 27 | M | Right | Right | 71-75 | 36 | 18 | 10 | Left basal ganglia stroke |
| 28 | M | Left | Left | 41-45 | 179 | 22 | 15 | Right intracerebral haemorrhagic stroke |
| 29 | M | Right | Right | 61-65 | 60 | 31 | 18 | Left basal ganglia haemorrhagic stroke |
| 30 | F | Right | Right | 36-40 | 16 | 28 | 8 | Left deep intracerebral haemorrhagic stroke |
| 31 | M | Right | Right | 76-80 | 30 | 25 | 5 | Left hemisphere stroke (unspecified) |
| 32 | M | Right | Right | 46-50 | 57 | 21 | 6 | Left striatocapsular stroke |
| 33 | M | Left | Right | 56-60 | 14 | 31 | No data | Left hemisphere stroke (unspecified) |
| 34 | F | Right | Right | 51-55 | 23 | 44 | 11 | Left thalamic stroke |
| 35 | M | Left | Right | 36-40 | 65 | 8 | 5 | Left hemisphere stroke (unspecified) |
| 36 | M | Right | Left | 81-85 | 21 | 18 | 11 | Right pontine haemorrhagic stroke |
| 37 | M | Right | Right | 66-70 | 14 | 30 | 9 | Left hemisphere stroke (unspecified) |
| 38 | M | Right | Left | 56-60 | 7 | 43 | 10 | Right MCA territory stroke |
| 39 | M | Right | Left | 26-30 | 18 | 50 | 18 | Right hemisphere stroke (unspecified) |
| 40 | M | Left | Left | 61-65 | 23 | 23 | No data | Right hemisphere stroke (unspecified) |

**Table S1. Clinical and demographic characteristics, along with lesion site, reported for each stroke patient.**

#### Selection of priors for HGF models

We selected prior values for our HGF models based on estimates obtained from our data through an ideal observer model. An ideal observer is defined as a model that adopts a range of parameter values aimed at minimising the overall surprise an agent experiences upon receiving a series of inputs (refer to Weber et al. 2020, for an example application of an ideal observer model). This approach was preferred over the use of previously reported prior values for the binary categorical HGF model, primarily because we used fewer trials than in previous experimental designs using this reversal learning task and using the HGF. Consequently, prior values used in previous studies may not be directly applicable to newer model implementations. We used the HGF release v7.1 in Matlab R2020b, and functions ‘tapas_ehgf_binary’.

Using ideal observer models, the group priors (means) on perceptual parameters ω2, ω3 were estimated using MATLAB function robustcov: [$\omega_2$, $\omega_3$] = [-3 0.6]. The prior variance on these parameters was 4, as in previous work (Diaconescu et al., 2014; Hein et al ., 2021). For complete details on our prior parameters on the winning model M3, see **Table S2**.

| Prior | Mean | Variance |
|---|---|---|
| $\kappa$ | log(1) | 0 |
| $\omega_2$ | -3 | 4 |
| $\omega_3$ | 0.6 | 4 |
| $\mu_2^{(0)}$ | 0 | 0 |
| $\sigma_2^{(0)}$ | log(0.1) | 0 |
| $\mu_3^{(0)}$ | log(1) | 1 |
| $\sigma_3^{(0)}$ | log(1) | 1 |

**Table S2. Priors (means and variances) on perceptual parameters and starting values of the belief distributions for the winning HGF model M3.** Quantities are estimated in their native space when they are unbounded, such as $\omega_2$, $\omega_3$. Conversely, quantities with a natural lower bound at zero, like $\kappa$, $\mu_3^{(0)}$, and $\sigma_3^{(0)}$, are estimated in log-space. In the winning HGF model M3, $\omega_2$, $\omega_3$, $\mu_3^{(0)}$, $\sigma_3^{(0)}$ were free parameters ($\kappa$, $\sigma_2^{(0)}$, $\mu_2^{(0)}$ were fixed). The prior variances are in the space in which the corresponding parameter is estimated. In models M1-M2, the inverse decision noise parameter ζ had the default prior of the HGF toolbox, with mean = log(48), and variance = 1.

### HGF supplementary equations

We used a three-level enhanced Hierarchical Gaussian Filter (eHGF) for binary inputs in our study, where the hidden states evolve as Gaussian random walks at levels 2 and 3, and the step size of each random walk depends on the variance at the next higher level (variance coupling). The update equations resulting from the approximate inversion of the three-level binary eHGF are given in Mathys et al. (2011, 2014) and Hess et al. (2025):

$$\mu_1^{(k)} = \mathrm{u}^{(k)}$$

$$\mu_2^{(k)} = \hat{\mu}_2^{(k)} + \kappa_1 \sigma_2^{(k)} \delta_1^{(k)}, \qquad \sigma_2^{(k)} = \frac{1}{1/\hat{\sigma}_2^{(k)} + \kappa_1^2 \hat{\sigma}_1^{(k)}}$$

$$\mu_3^{(k)} = \hat{\mu}_3^{(k)} + \sigma_3^{(k)} \frac{\kappa_2}{2} \hat{w}_2^{(k)} \hat{\delta}_2^{(k)}, \quad \pi_3^{(k)} = \hat{\pi}_3^{(k)} + \max\left\{0, \frac{\kappa_2^2}{2} w_2^{(k)} \left(w_2^{(k)} + r_2^{(k)} \delta_2^{(k)}\right)\right\}$$

These equations make use of the following definitions:

$$\delta_1^{(k)} \stackrel{\text{def}}{=} \mu_1^{(k)} - \hat{\mu}_1^{(k)}$$

$$\hat{\mu}_1^{(k)} \stackrel{\text{def}}{=} s\left(\kappa_1 \hat{\mu}_2^{(k)}\right)$$

$$\hat{\sigma}_1^{(k)} \stackrel{\text{def}}{=} \hat{\mu}_1^{(k)}\left(1 - \hat{\mu}_1^{(k)}\right)$$

$$\hat{\mu}_2^{(k)} = \mu_2^{(k-1)} + t^{(k)}\rho_2$$

$$\hat{\sigma}_2^{(k)} \stackrel{\text{def}}{=} \sigma_2^{(k-1)} + e^{\left(\kappa_2 \mu_3^{(k-1)} + \omega_2\right)}$$

$$\hat{\mu}_3^{(k)} = \mu_3^{(k-1)} + t^{(k)}\rho_3$$

$$\pi_3^{(k)} \stackrel{\text{def}}{=} \frac{1}{\sigma_3^{(k)}}$$

$$\hat{\pi}_3^{(k)} \stackrel{\text{def}}{=} \frac{1}{\sigma_3^{(k-1)} + t^{(k)} e^{(\omega_3)}}$$

$$\hat{w}_2^{(k)} \stackrel{\text{def}}{=} \frac{t^{(k)} e^{\left(\kappa_2 \mu_3^{(k-1)} + \omega_2\right)}}{\sigma_2^{(k-1)} + e^{\left(\kappa_2 \mu_3^{(k-1)} + \omega_2\right)}}$$

$$w_2^{(k)} \stackrel{\text{def}}{=} \frac{t^{(k)} e^{\left(\kappa_2 \mu_3^{(k)} + \omega_2\right)}}{\sigma_2^{(k-1)} + t^{(k)} e^{\left(\kappa_2 \mu_3^{(k)} + \omega_2\right)}}$$

$$r_2^{(k)} \stackrel{\text{def}}{=} \frac{t^{(k)} e^{\left(\kappa_2 \mu_3^{(k)} + \omega_2\right)} - \sigma_2^{(k-1)}}{\sigma_2^{(k-1)} + t^{(k)} e^{\left(\kappa_2 \mu_3^{(k)} + \omega_2\right)}}$$

$$\hat{\delta}_2^{(k)} \stackrel{\text{def}}{=} \frac{\sigma_2^{(k)} + \left(\mu_2^{(k)} - \mu_2^{(k-1)}\right)^2}{\sigma_2^{(k-1)} + e^{\left(\kappa_2 \mu_3^{(k-1)} + \omega_2\right)}} - 1$$

$$\delta_2^{(k)} \stackrel{\text{def}}{=} \frac{\sigma_2^{(k)} + \left(\mu_2^{(k)} - \mu_2^{(k-1)}\right)^2}{\sigma_2^{(k-1)} + t^{(k)} e^{\left(\kappa_2 \mu_3^{(k)} + \omega_2\right)}} - 1$$

In these equations, $u^{(k)}$ denotes the binary input on trial $k$, $x_i^{(k)}$ are the hidden states, and $\kappa_2$, $\omega_2$, and $\omega_3$ are parameters governing their evolution. Temporal drift can be modulated by parameters $\rho_2$ and $\rho_3$, while $\kappa_1$ scales the coupling from level 2 to level 1. In our application, we set $\kappa_1$=1, $\rho_2$= $\rho_3$=0, and $t^{(k)}$=1 for all trials, effectively removing their influence. Accordingly, in the main manuscript, when we write $\kappa$, we refer to $\kappa_2$, which governs the coupling between levels 2 and 3.

### HGF Simulations

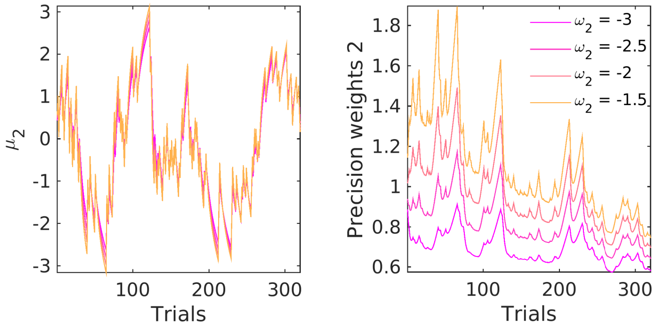


**Figure S1.** Simulated belief trajectories and precision weight terms on level 2. **Left**: In the winning model, M3, belief trajectories for $\mu_2$ were simulated using fixed input data, with priors set at $\mu_2^{(0)} = 0$, $\sigma_2^{(0)} = 0.1$, $\mu_3^{(0)} = 1$, $\sigma_3^{(0)} = 1$, $\kappa = 1$, $\omega_3 = -0.5$, but with $\omega_2$ being modulated. This parameter, $\omega_2$, represents the tonic part of the variance in the Gaussian random walk for $x_2$ and modulates the learning rate about response outcomes at the lowest level. The graphic demonstrates that decreasing $\omega_2$ (represented by changes from orange to magenta lines) is associated with smaller update steps in the belief about the action-outcome contingency tendency, $\mu_2$. **Right**: In line with the left panel, decreasing $\omega_2$ also reduces the estimation uncertainty about the reward tendency, $\sigma_2$. Since $\sigma_2$ represents the precision weights term scaling belief updating at level 2, this simulation indicates that lower tonic volatility, $\omega_2$, has a slowing effect on belief updating about the action-outcome contingency tendency. Image adapted with permission from reference[34].

### HGF with multimodal response models

To consider an alternative motor vigour analysis approach, we assessed a recently developed extension of the eHGF that incorporates a set of response models enabling simultaneous inference from categorical (binary) choices and continuous reaction times (RTs; Hess et al., 2025). This modelling framework has been validated through simulations and empirical data from healthy participants completing a speed-incentivised associative reward learning (SPIRL) task, which includes binary decision-making in a volatile environment, as in our one-armed bandit task, and speeded RTs.

Within this framework, in addition to estimating individual decision-making parameters via hierarchical Bayesian inference, researchers can also estimate the regression coefficients describing the association between trial-wise computational variables of interest and log-transformed RT (*logRT*), as the models include a general linear model (GLM) of *logRT* as the dependent variable and several belief trajectories as eHGF regressors.

Accordingly, this approach provides a powerful means of jointly assessing belief updating during decision-making and motor vigour. However, it has not yet been validated in samples differing in baseline RTs, such as individuals with Parkinson's disease or stroke, who are known to exhibit slower responses. We therefore ran some analyses to evaluate the suitability of this approach for inferring decision-making parameters under different absolute RT values.

First, we used pilot data from Hess et al. (2025), consisting of 20 healthy participants who completed their task, and fitted the winning model from that study (model M1), identified as the best-fitting model in a larger sample of 59 right-handed healthy individuals. This model comprised the perceptual eHGF and two response models: the standard unit-square sigmoid for binary categorical responses, and the following observation model for continuous *logRT* responses:

$$\log(\mathrm{y}_{\mathrm{rt}}^{(\mathrm{k})}) \sim \mathcal{N}\left(\beta_0 + \beta_1 \mathrm{S}^{(k-1)} + \beta_2 \hat{\sigma}_1^{(k)} + \beta_3 \hat{\sigma}_2^{(k)} + \beta_4 \exp^{\hat{\mu}_3^{(k)}}\right)$$

with

$$S^{(k-1)} \stackrel{\mathrm{def}}{=} \begin{cases} -\log_2(\hat{\mu}_1^{(k-1)}) & \text{if } u^{(k-1)} = 1, \\ -\log_2(1-\hat{\mu}_1^{(k-1)}) & \text{if } u^{(k-1)} = 0 \, . \end{cases}$$

The regressors in the GLM above included several belief trajectories from the perceptual model, such as the agent's estimated uncertainty about the outcome ($\hat{\sigma}_1^{(k)}$) on trial $k$, and the expected volatility on trial $k$ (exponential of expected log-volatility $\hat{\mu}_3^{(k)}$). Interestingly, in Hess et al. (2025), only the regression coefficient $\beta_2$ was significantly different from zero (and positive) across participants, suggesting that logRT in this task was primarily modulated by trial-wise estimated outcome uncertainty, $\hat{\sigma}_1^{(k)}$.

To fit model M1 to the pilot data from Hess et al. (2025), we used the code provided by the authors (available at https://gitlab.ethz.ch/tnu/code/hessetal_spirl_analysis). We also used the same versions of the open-source software packages reported in their publication: the HGF Toolbox (v7.1) within the 'Translational Algorithms for Psychiatry-Advancing Science' (TAPAS v6.0.1, commit 604c568) package (Frässle et al., 2021), and the Variational Bayesian Analysis Toolbox (VBA, commit aa46573; Daunizeau et al., 2014). Following the authors' recommendations in the main analysis script (main_local_serial.m), we set local_cores to match the number of cores on our local machine and reduced the number of random initialisations of the optimisation algorithm during model inversion by defining options.opt_config.nRandInit = 0 in the spirl_specs.m function.

As in Hess et al. (2025), the prior eHGF parameters used to fit the pilot data were the default values provided in the HGF Toolbox. The prior on *logRT* for the continuous-response model is the average *logRT* across trials and participants: 5.91 log-ms.

Fitting model M1 to the pilot dataset of 20 participants (sample 1) yielded individual perceptual model parameters for the eHGF and the associated regression coefficients for the continuous-response model in each participant. The trial-average *logRT* values across participants ranged from 5.50 log-ms (≈ 245 ms) to 6.34 log-ms (≈ 567 ms).

Next, we selected a random subset of 10 participants from this sample and increased their individual *logRT* values by a constant offset of 0.2 log-ms, while preserving their trial-wise variation. Thus, if a participant's responses became faster or slower across trials as a function of $\hat{\sigma}_1^{(k)}$, the same pattern was retained, but superimposed on an overall slower baseline *logRT*. The remaining 10 participants retained their original *logRT* values, effectively creating one group of slower-RT participants (analogous to an experimental group) and one group with normative Rts. See the timecourse of *logRT* for example participants with slower RTs (IDs 1, 4, 20) in **Figure S2a-c.**

In this modified dataset (sample 2), the prior on *logRT* for the continuous-response model was updated to the new grand mean across trials and participants (6.00 log-ms).

Fitting M1 under this modified sample, we observed that key parameters of the perceptual model changed, despite participants categorical decision making behaviour being identical (**Figure S2d-f**).

We next asked whether this unexpected model behaviour could be attributed to changes in the prior on *logRT*. To examine this, we created a new sample (sample 3) from the pilot data in which 10 participants had an increased baseline shift in *logRT* of +0.2 log-ms, while the remaining 10 participants had a corresponding decrease of –0.2 log-ms. This manipulation effectively maintained the same mean prior on *logRT* for the continuous-response model (5.91 log-ms). Nevertheless, key parameters of the perceptual model again changed, even though participants' categorical decision-making behaviour was identical and the prior on *logRT* remained unchanged (**Figure S2d-f**).

Comparing the belief trajectories for these example participants and samples, we observe that whether a participant exhibits slower or faster belief updating (dynamics of $\mu_2$ and $\mu_3$) is modulated by their baseline logRT. Participant #1, for instance, showed slower belief updating in $\mu_2$ under sample 3 compared with faster updating in samples 1 and 2, which was associated with a lower $\omega_2$, as expected. Similar model behaviour was observed for participant #20. In contrast, participant #4, who also experienced changes in baseline logRT across samples 2 and 3, displayed essentially identical eHGF parameters and belief trajectories across samples. Correspondingly, the log-model evidence (LME) changed from sample 1 to samples 2 and 3 for participants #1 and #20, but not for participant #4.

Panels **Figure S2g-i** summarise the group-level distributions (boxplots) of key model parameters, including the GLM regression coefficient $\beta_2$ for the outcome uncertainty regressor $\hat{\sigma}_1$ (j), $\omega_2$ (k), and $\omega_3$ (l). These panels illustrate that the distribution of model parameters differs unexpectedly across samples, both for the regression coefficient $\beta_2$, which is part of the observation model for continuous RT responses (GLM), and for the perceptual parameters of the eHGF.

Accordingly, our analyses indicate that, for participants exhibiting identical decision-making behaviour, this modelling approach infers different perceptual learning dynamics depending on their absolute *logRT*.

This interesting behaviour of the new eHGF framework for multimodal response models suggests that further work is required to fully understand these effects, and that additional validation is needed in participant samples with markedly different RTs, such as individuals with Parkinson's disease or stroke, before this model can be reliably deployed in clinical settings where RTs are substantially slower than in a healthy control sample.

In summary, the motor-vigour analyses reported in the main manuscript followed our established approach: fitting separate eHGF perceptual models to explain belief updating during learning, and using Bayesian mixed models to separately assess motor vigour.

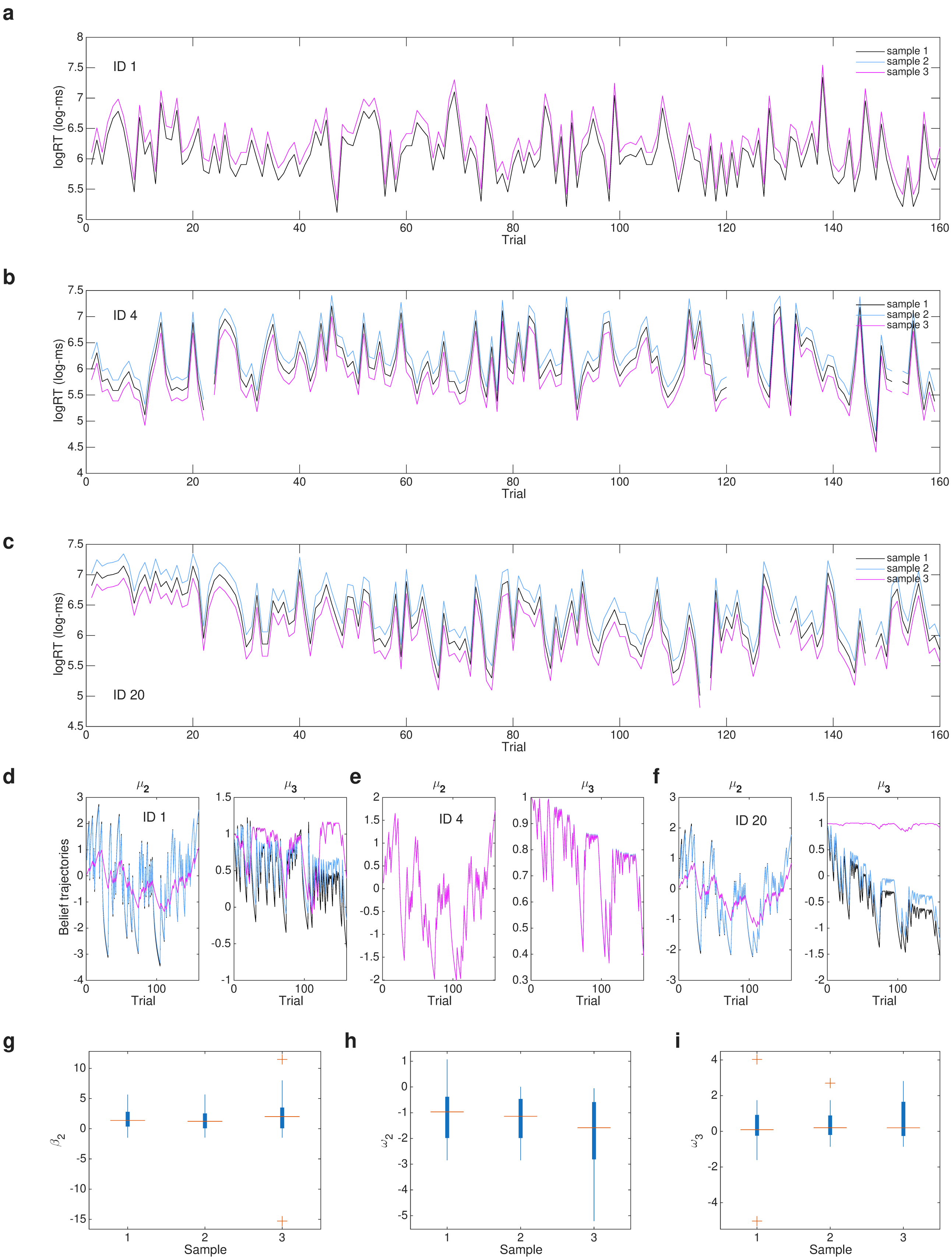


**Figure S2. Individual and group-level model results. (a–c)** Trial-by-trial log response times (*logRT*, in log-ms) for three selected participants (IDs 1, 4, 20) and for three samples derived from the 20 pilot participants from Hess et al (2025):

sample 1 (black line), original data; sample 2 (blue line), a random subset of 10 participants with an increase in baseline *logRT* of +0.2 log-ms (trial-wise variation preserved), while the remaining 10 participants retained their original values; sample 3 (magenta line), a random subset of 10 participants with an increase of 0.2 log-ms and the remaining 10 with a decrease of -0.2 log-ms in baseline *logRT*. **(d–f)** Belief trajectories for the same three participants, showing inferred posterior means of level-2 beliefs ($\mu_2$; left) and level-3 beliefs ($\mu_3$; right) from the perceptual model eHGF under M1. **(g–i)** Group-level distributions (boxplots) of relevant model parameters, including the GLM regression coefficient $\beta_2$ for the outcome uncertainty regressor $\hat{\sigma}_1$ (j), $\omega_2$ (k), and $\omega_3$ (l).

#### Parameter estimation in the HGF

Back to the HGF analysis in the main manuscript, we conducted simulations to evaluate the reliability of parameter estimates in our implementation of the best-fitting eHGF model (M3). In this model, the free parameters estimated for each individual were $\omega_2$, $\omega_3$, $\mu_3^{(0)}$ and $\sigma_3^{(0)}$ , with priors as defined in (**Table S2**).

We simulated behavioural responses from 100 agents for six different values of $\omega_2$ (600 simulations in total) and five different values of $\omega_3$ (500 simulations in total), each using the input sequences of two different participants (one SK and one HC). To assess the accuracy of estimating $\mu_3^{(0)}$, we run additional simulations with 100 agents for six different values of $\mu_3^{(0)}$, and six values of $\sigma_3^{(0)}$.

All simulations were implemented using the function tapas_simModel.m from the HGF toolbox, iterating over $\omega_2$, $\omega_3$ (denoted as om2 and om3, respectively) and over the number of iterations (N = 100). The priors on free parameters were defined as in **Table S2.**

```
sim = tapas_simModel(u, 'tapas_ehgfsim_binary',  [NaN 0 1 NaN 1 1 NaN 0 0 1 1 NaN om2 om3],
'tapas_unitsq_sgm_mu3',123456789);
```

Here, function tapas_ehgfsim_binary.m is a copy of the default HGF function tapas_ehgf_binary.m but with priors on $\mu_3^{(0)}$ and $\sigma_3^{(0)}$ as used in our study (**Table S2**). Simulated responses were then fit using:

The simulated behavioural responses *sim.y* and the input *u* observed by each example participant were then fitted with the tapas_fitModel.m function, similarly to the way we fitted the standard empirical data in our participants:

```
tapas_fitModel(sim.y, u, ehgf_binary_config, unitsq_sgm_mu3_config, optim_config)
```

with

```
optim_config = tapas_quasinewton_optim_config()
unitsq_sgm_mu3_config = tapas_unitsq_sgm_mu3_config()
```

```
ehgf_binary_config = tapas_ehgf_binary_config()
```

This analysis demonstrated high accuracy in estimating $\omega_2$ and $\mu_3^{(0)}$, whereas $\omega_3$ was recovered with lower accuracy and $\sigma_3^{(0)}$ was poorly recovered, consistent with previous work (Hein et al., 2021; Reed et al., 2020; see main text). See figure below:

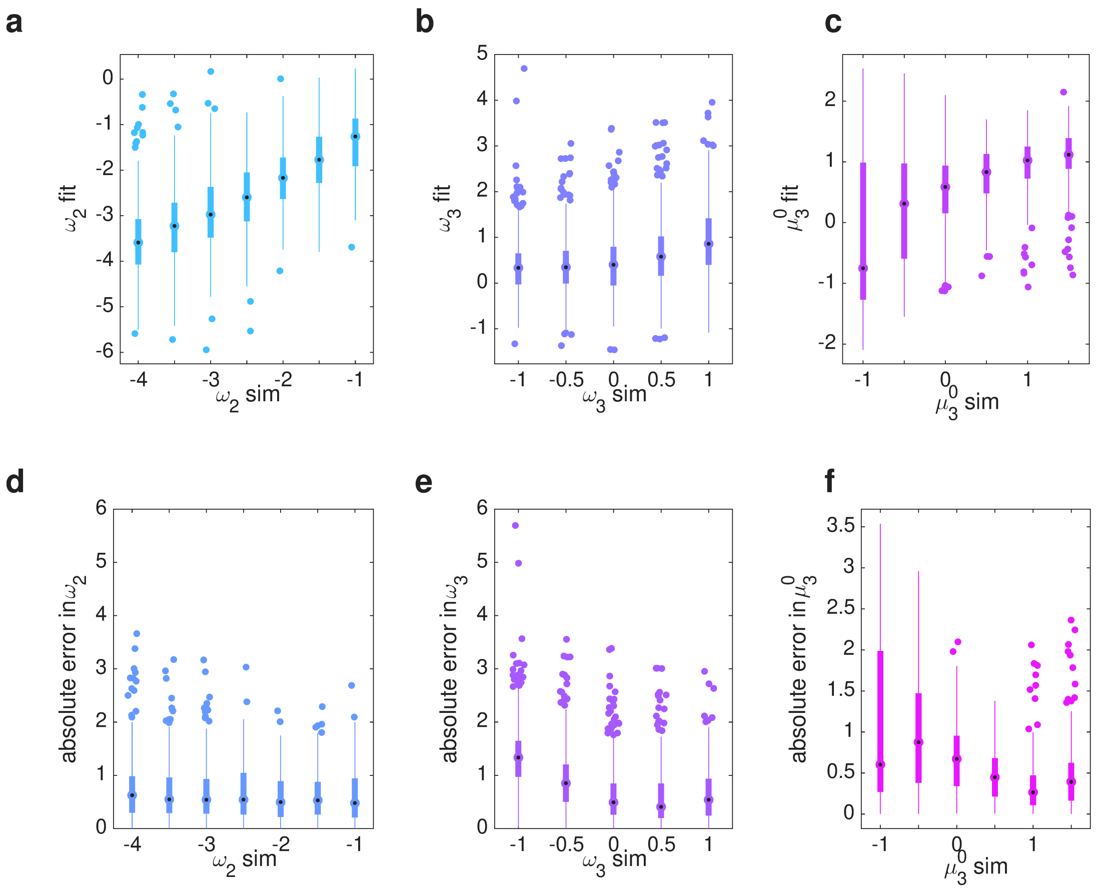


**Figure S3. HGF parameter estimation using the input observed by one participant (ID 63, healthy control). a-b)** Boxplots (median, 25 and 75 percentiles) illustrating the results of parameter estimation for $\omega_2$ (a), $\omega_3$ (b), and $\mu_3^{(0)}$ (c). The x-axis represents the set parameters introduced in the simulated responses (labelled “sim”), while y-axis data reveal the corresponding estimated value of that same parameter (labelled “fit”). Parameters $\omega_2$ and $\mu_3^{(0)}$ were estimated with high accuracy, as there was a high significant correlation between simulated and estimated (fit) values: Pearson R = 0.667, $P \ll 1 \times 10^{-6}$ for $\omega_2$, R = 0.508, $P \ll 1 \times 10^{-6}$ for $\mu_3^{(0)}$. Parameter $\omega_3$ was estimated less accurately, though the association between simulated and fitted values was still significant: R = 0.2501, $P \ll 1 \times 10^{-6}$. By contrast, $\sigma_3^{(0)}$ showed poor recovery, R = 0.016, $P$ = 0.507. Similar parameter estimation results were obtained in separate simulations for an agent observing the input from one stroke participant (ID 2): Pearson R = 0.7430, $P \ll 1 \times 10^{-6}$ for $\omega_2$, R = 0.453, $P \ll 1 \times 10^{-6}$ for $\mu_3^{(0)}$, R = 0.2331, $P \ll 1 \times 10^{-6}$ for $\omega_3$, R = 0.017, $P$ = 0.473 for $\sigma_3^{(0)}$.

### Motor vigour analyses

| Model # | Model |
| --- | --- |
| 1 | y ~ 1 + prediction.c |
| 2 | y ~ 1 + group*prediction.c |
| 3 | y ~ 1 + group*prediction.c*arm |
| 4 | y ~ 1 + prediction.c + (1\|subject) |
| 5 | y ~ 1+ group*prediction.c + (1\|subject) |
| 6 | y ~ 1 + group*prediction.c*arm + (1\|subject) |
| 7 | y ~ 1+ prediction.c+(1+prediction.c\|subject) |
| 8 | y ~ 1+ group*prediction.c + (1+prediction.c\|subject) |
| 9 | y ~ 1+ group*prediction.c*arm + (1+prediction.c\|subject) |
| | The brms family was Gaussian |

**Table S3.** Bayesian Multilevel Models with a Gaussian distribution assessing the effect of strength of predictions about action-reward contingencies on movement time (logMT). Models of increasing complexity were defined to assess whether logMT (represented by y) was modulated by the strength of predictions about action-outcome contingencies $|\hat{\mu}_2|$. This continuous predictor was centred and is denoted by *prediction.c* in the table. Factors group and arm were categorical variables. The most complex model, M9, included a three-way interaction between group, arm, and *prediction*.c, and by-subject random effects on both the intercept and the slope of the relationship between movement time and uncertainty. Similar models were constructed for the predicted uncertainty about outcomes, $\hat{\sigma}_1$ (*uncertainty.c*).

Additional models 1–9 replaced the *prediction.c* regressor with trial-wise informational uncertainty at the outcome level (Bernoulli variance $\hat{\sigma}_1$, centered as *uncertainty.c*), which has also been shown to explain the relationship between decision-making and movement time (see main text).

Model comparison across all 18 models (1–9 for *prediction.c*, and 1–9 for *uncertainty.c*) indicated that the performance of each model structure was comparable for the two regressors. Model 9 with the *prediction*.c regressor emerged as the best-fitting model, yet performing equivalently to the version including *uncertainty*.c, as indicated by LOO-CV (elpd_diff = -0.7860, not exceeding 2 x se_diff = 2 x 2.7908). Both outperformed the next-best alternative (model 8; elpd_diff = -182.95, > 2 x se_diff = 2 x 19.21). Given that ELPD differences greater than four with sufficient observations (>100) support reliable model comparison via standard error estimation (Sivula et al., 2020), these results indicate a substantial improvement of model 9 over alternatives. However, both model 9 variants were equivalent. The regression coefficients for the uncertainty.c variant of model 9 are reported in **Table S4.**

| **Model 9 Dependent Variable** | **Parameter (Population-level effects)** | **Estimate** | **l-95% CrI** | **u-95% CrI** | **R-hat** |
|---|---|---|---|---|---|
| Movement time (logMT, in log-ms) | | | | | |
| | Intercept | **6.715** | **6.629** | **6.803** | 1.00 |
| | group (SK-HC) | **0.231** | **0.121** | **0.342** | 1.00 |
| | uncertainty | **0.387** | **0.011** | **0.771** | 1.00 |
| | limb (weak - strong) | **-0.073** | **-0.093** | **-0.052** | 1.00 |
| | group:uncertainty | 0.258 | -0.248 | 0.749 | 1.00 |
| | group:limb | **0.288** | **0.260** | **0.315** | 1.00 |
| | uncertainty:limb | -0.290 | -0.605 | 0.018 | 1.00 |
| | group:uncertainty:limb | -0.220 | -0.638 | 0.204 | 1.00 |

**Table S4.** Summary parameter estimates for the best-fit Bayesian multilevel model assessing the effect of uncertainty about the outcome on timing performance, which was comparable to model 9 for *prediction.c* (based on LOO-CV). Estimate = posterior mean; CrI = credible interval based on quantiles. Gelman-Rubin statistics demonstrate excellent chain convergence (R-hat < 1.01). The effective sample size (ESS) was >> 400 for each parameter estimate, denoting good convergence. The predictor "uncertainty" denotes the centred values of the informational uncertainty about the outcome ($\hat{\sigma}_1^{(k)}$), *uncertainty.c*. Credible effects are considered when the 95% lower and upper-bound CrI around the Bayesian point estimate do not include zero (denoted by bold font). Among several effects, this model showed a credible effect of greater trial-wise uncertainty being associated with slower MTs, indicated by a positive coefficient for *uncertainty.c* (posterior estimate = 0.387, 95% CrI [0.011, 0.771]), with the CrI excluding zero.

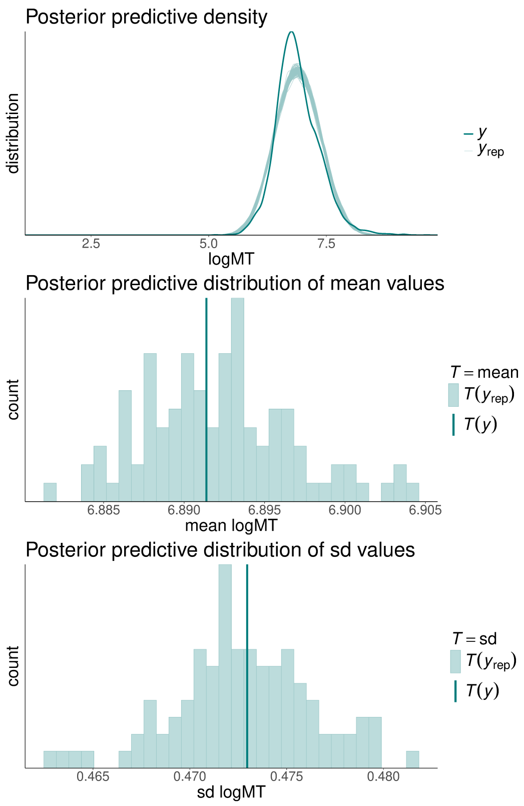

**Figure S4. Posterior predictive checks for Model M9. Top**: Posterior predictive density plot comparing the empirical distribution (dark green) with 100 datasets simulated from the posterior (light green). **Middle–Bottom**: Posterior predictive distributions of summary statistics. The middle panel shows the distribution of simulated means; the bottom panel shows simulated standard deviations. Vertical lines indicate empirical values. Priors for M9 were specified as described in the main text.
